# Identification and Reproducibility of Novel Combinatorial Genetic Risk Factors for Endometriosis across UK and US Patient Cohorts

**DOI:** 10.1101/2025.08.13.25333595

**Authors:** JM Sardell, S Das, GL Møller, M Sanna, K Chocian, K Taylor, AR Malinowski, C Stubberfield, A Rochlin, S Gardner

## Abstract

**Background:** Although endometriosis affects approximately 10% of women, timely diagnosis and effective treatments remain out of reach for many patients. Relatively little is known about the genetic drivers of endometriosis even though its heritability is around 50%. Two recent large genome wide association study meta-analyses (meta-GWASs) have identified up to 82 genomic loci associated with risk of endometriosis, but together these explain a small proportion of disease risk.

**Methods:** We used the PrecisionLife® combinatorial analytics platform to identify disease signatures (i.e., unique combinations of 2-5 SNPs) that were significantly associated with endometriosis in a White British UK Biobank (UKB) cohort. We assessed the reproducibility of these disease signatures, as well 35 of the 42 previously reported 2023 meta-GWAS SNPs, in a multi-ancestry US endometriosis cohort from All of Us (AoU) after controlling for population structure. To ensure that the results do not disfavor historically underserved patient populations, we explicitly evaluated the reproducibility of these signatures across patient groups with different self-reported race/ethnicities. Finally, we characterized drug repurposing opportunities for a subset of the novel candidate genes.

**Results:** We identified 1,709 disease signatures, comprising 2,957 unique SNPs, that were significantly associated with increased prevalence of endometriosis in the White British UKB cohort. Reproducibility rates in all AoU participants were significantly higher than random and highest for higher frequency disease signatures, ranging from over 68% for signatures with greater than 4% frequency up to 80-88% for signatures with greater than 9% frequency. Reproducibility rates remained broadly unchanged for most frequency bins within self-identified Black/African American and Hispanic/Latino AoU sub-cohorts. Pathways enriched in reproducible disease signatures included cell adhesion, proliferation and migration, cytoskeleton remodeling, and angiogenesis as well as biological processes involved in fibrosis and neuropathic pain.

39% of signatures (667 / 1709) mapped to at least one gene previously linked to endometriosis via the meta-GWAS studies, including 20% of signatures (336 / 1709) that contained at least one meta-GWAS lead SNP. The remaining disease signatures (1042 / 1709) were completely novel. Several novel signatures contained SNPs that mapped to genes that were previously associated with endometriosis but not identified by the meta-GWAS.

We identified 79 entirely novel candidate genes, of which 33 are tractable drug targets. Of the 25 top SNPs ranked based on frequency in signatures, we identified 9 genes that primarily occurred in entirely novel signatures, all of which demonstrated high rates of reproducibility. These 9 genes provide support for hypothesized links between endometriosis and autophagy and macrophage biology.

**Conclusions:** This study identified a large number of reproducible novel disease signatures, SNPs, and novel candidate genes associated with increased prevalence of endometriosis. It provides support for prior findings from meta-GWAS and other studies linking specific genes to endometriosis. These findings suggest the potential to inform new therapeutic or targeted drug repurposing/repositioning programs aimed at development of new and better treatment options for patients.

**Lay Sunnary:** Endometriosis affects about one in 10 women, usually between the ages of 15 and 49. It is difficult to diagnose and hard to treat as there are very few effective drugs available. Previous research has found some genes linked to the disease, but we wanted to find more so that better new drugs can be designed that will help more patients. By using a new way to study the disease, we found 79 more genes linked to endometriosis. We believe many of these could be targeted either by new drugs or by existing drugs that were originally developed for another disease. We also showed that these are likely to work for people from different communities, so they could benefit as many patients as possible.

**Graphical Abstract:** Graphical abstract.
A. Discovery of novel combinatorial genetic associations with endometriosis in a White British patient population from UK Biobank (UKB). B. Analysis of the reproducibility of the UKB disease signatures and 35 of the 42 SNPs identified by the 2023 meta-GWAS analysis in a mixed ancestry US population from All of Us and identification of 79 novel, high frequency candidate genes associated with endometriosis in both populations.

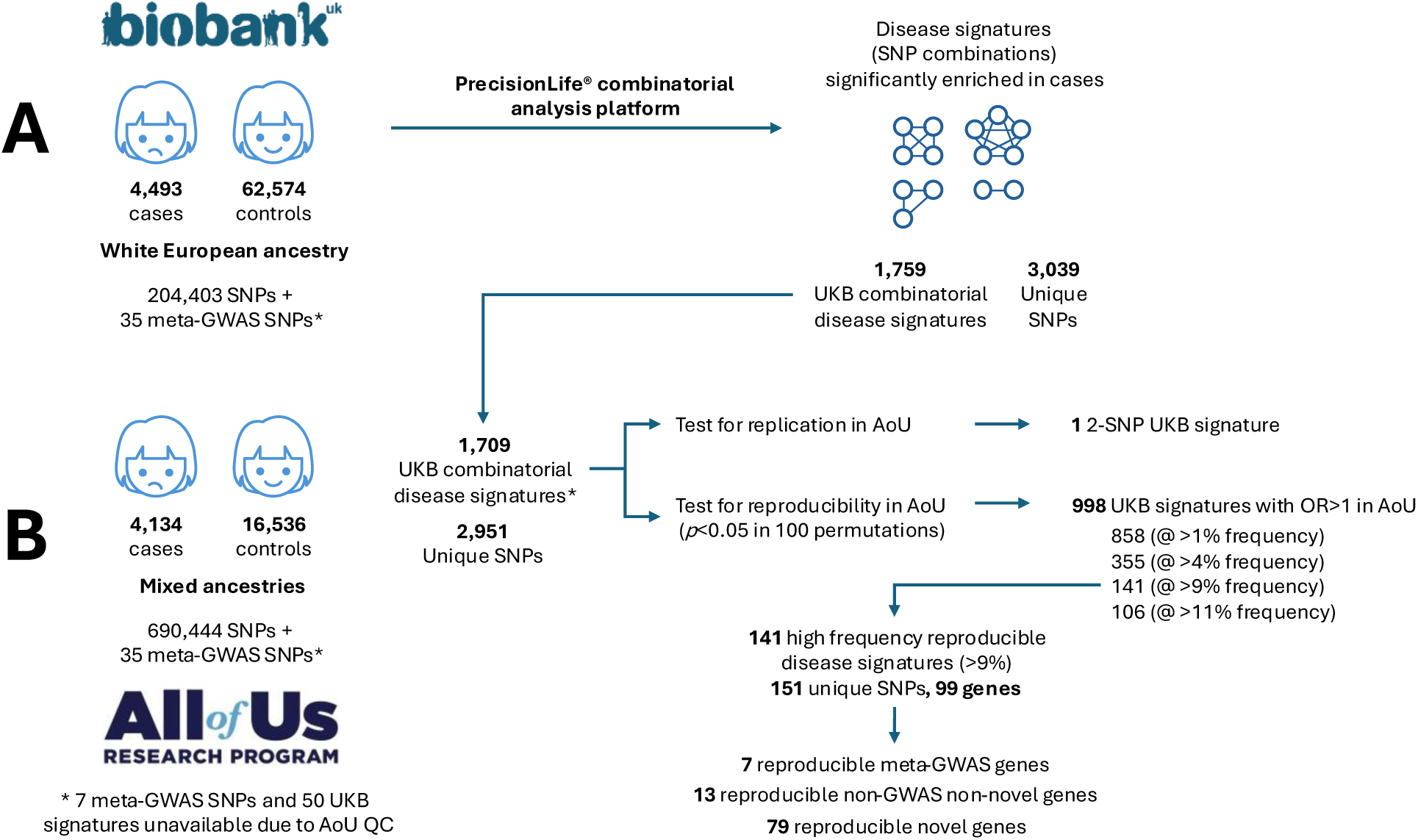

## Introduction

Endometriosis is a debilitating and, in many cases, progressive chronic disease that impacts approximately 10% of reproductive age women (1,2). Despite its high prevalence and severe pathophysiology, the average time to diagnosis of endometriosis from onset of symptoms is almost nine years in the UK, and treatments such as invasive surgery and hormonal therapies often have limited symptomatic impact (3,4) and significant side effects (5,6).

The long delay between onset of symptoms, diagnosis, and effective treatment occurs in part because the mechanisms underlying the development of endometriosis and its associated symptoms and comorbidities, such as chronic pain and infertility, are poorly understood (7). Several hypotheses have been proposed to explain the development of endometriosis, including retrograde menstruation, alterations in the peritoneal fluid, and bacterial infection. However many of these have contradicting evidence, and none are definitive (8).

A major challenge for research is that endometriosis is a multi-factorial disease, believed to be caused by complex interactions between genetic, hormonal, and environmental factors (9). Two recent genome-wide association study (GWAS) meta-analyses have together identified up to 82 risk loci for endometriosis (10,11). The larger 2026 study analyzed 105,869 endometriosis cases and 1,282,731 controls across multiple ancestries from eight cohorts including UK Biobank and All of Us. It identified 80 loci, of which 37 were novel, implicated in cell differentiation, immune and hormonal regulation, tissue remodeling and inflammation (10). The 2023 study was based on 60,674 endometriosis cases and 701,926 controls of predominantly European ancestry (98%) from 24 cohorts including UK Biobank (but not All of Us), and identified 42 loci, several mapping to genes linked to pain perception and maintenance (11).

These loci nonetheless explain only a small proportion of disease risk. In the 2026 study, SNP-based heritability was estimated at 6% in European cohorts while the genome-wide significant signals in the 2023 study explained at most 5% of disease variance, indicating that much remains unknown about the biology of this complex disease (10,11).

Many affected women schedule ten or more medical consultations before receiving an endometriosis diagnosis (12). The current gold standard for diagnosis is surgical confirmation of the presence of endometrial lesions (13), but these surgeries are invasive, painful and carry a degree of risk (14). While the sensitivity and accuracy of non-invasive imaging methods for diagnosing the disease are improving, they continue to vary enormously depending on the technique, type, size, and location of lesions, and experience of the operator (15). Thus, there is pressing need for more effective treatment options and more accurate, ideally non-invasive, diagnostic tools for endometriosis.

### Conbinatorial Analytics for Conplex Diseases

Combinatorial analysis enables hypothesis-free identification of combinations of genetic variants (‘disease signatures’) that are significantly over – or under-enriched in patients with a disease or other specific phenotype. The disease signatures, which are comprised of between 1 to 5 SNP-genotypes, capture both the linear and non-linear (e.g., epistatic) interactions between multiple genomic loci that reflect the complex disease biology involved in heterogeneous chronic diseases. By capturing this additional signal, combinatorial disease risk signatures expand our understanding of complex diseases beyond the single SNP associations identified by GWAS (16).

The PrecisionLife® combinatorial analytics platform has been used in multiple studies to identify key genes and genetic mechanisms associated with disease risk and resilience including amyotrophic lateral sclerosis (ALS) (17), myalgic encephalomyelitis / chronic fatigue syndrome (ME/CFS)(18,19), and long COVID (20). These studies have demonstrated that, by combining additive and non-additive signals from multiple genetic variants, combinatorial analysis can identify significant novel genetic associations in smaller datasets than are required for GWAS. Validation of the approach has been achieved via studies of reproducibility and replication in independent cohorts, both by the authors (19,21) and independent researchers (22).

### Ains of Study

We hypothesized that combinatorial analysis would identify reproducible genetic signals for endometriosis that complement and extend previous findings from GWAS and other studies. At the time this study was designed and analyzed, only the 2023 GWAS meta-analysis was available.

We first ran combinatorial analysis on a cohort of endometriosis patients and controls from a UK Biobank (UKB) dataset. This analysis identified 1,709 disease signatures, each comprising between 2 to 5 SNP-genotypes, that were significantly associated with increased risk of endometriosis.

We then sought to demonstrate significant enrichment of reproducing disease associations for these signatures in an independent, diverse ancestry endometriosis cohort from All of Us (AoU) for which we had overlapping genotype data. We wanted to evaluate the relative reproducibility of the disease signatures across all participants and in sub-cohorts of self-reported Black/African American and Hispanic/Latino patients to ensure that the findings of this analysis are unlikely to exacerbate inequalities in medical treatment for historically underserved populations.

To demonstrate the utility of the combinatorial analytics approach, we sought to establish reproducibility of disease signatures that were not linked to the 82 significant meta-GWAS loci, which identify unambiguously novel genetic disease associations with endometriosis. We annotated these novel candidate genes from a biological perspective to inform the broader understanding of the biological drivers of endometriosis and used our knowledge of the modulators of these novel candidate genes (with appropriate directionality to identify potential drug repurposing candidates for the associated mechanisms)(23).

## Materials and Methods

### Generation of the UK Biobank Endonetriosis Cohort

We used a cohort of endometriosis patients and healthy controls from UKB to identify multi-SNP disease signatures that are significantly associated with elevated prevalence of endometriosis in a case-control design. Cases were initially defined as any UKB participant with self-reported diagnosis (Data field 20002: data-code 1402) or ICD-10/9 codes (N80.*) of endometriosis as of April 2021. We further removed all patients with adenomyosis (ICD-10 code N80.0 – ‘endometriosis of uterus’).

To minimize the risk of detecting false positive disease signatures that are indirectly associated with disease due to population substructure rather than disease biology, we filtered the cohort to include only UKB participants identified as having self-reported ‘White British’ genetic ancestry. We were unable to perform combinatorial analyses on UKB patients with alternative ancestries due to their low representation and small sample sizes in that dataset.

196,188 potential controls were identified that matched the following criteria:

- Females with self-reported White British genetic ancestry
- Excluding all patients with self-reported diagnosis (Data field 20002: data-code 1402) or ICD-10/9 codes (N80.*) for endometriosis

We then randomly selected a subset of controls to produce a 1:10 case-control ratio relative to the number of UKB patients with all forms of endometriosis (N80.*).

The combinatorial analysis study was hypothesis-free. However, alongside the signatures identified by combinatorial analysis, we also wished to test the relative reproducibility of the lead SNPs identified by the 2023 meta-GWAS study (11), which was available at the time of this study’s initiation (hereafter referred to as ‘2023 meta-GWAS SNPs’). Of the 42 SNPs reported by the 2023 meta-GWAS, only 7 were present on the genotyping array used by UKB. To allow us to capture the effects of these missing loci in our combinatorial analysis, we incorporated genotypes for the missing meta-GWAS SNPs using imputed SNP genotype data provided by UKB.

We would not typically conduct combinatorial analysis on imputed genotype data as the systematic genotyping errors associated with imputation compound when considering combinations of SNP genotypes, especially across ancestries. For example, a 2% average misgenotyping error rate for individual SNPs implies that nearly 10% of samples will likely be misgenotyped for at least one of the SNPs in a five-SNP disease signature.

However, it was considered that the UKB imputation error rate is lowest for White British samples, and that inclusion of 42 imputed meta-GWAS SNPs was unlikely to result in compounded error in this study as they comprise a very small fraction (0.02%) of the total dataset. Indeed, we identified only 9 disease signatures (out of 1,709) that contained more than one meta-GWAS SNPs, and no disease signature contained more than two meta-GWAS SNPs, suggesting minimal effects of compounding imputation error.

In any combinatorial analysis, only a small fraction of the total possible number of disease signatures can be evaluated due to computational and statistical power limitations, and it would be inefficient to devote limited resources to evaluating signatures that could not be further validated using the available datasets. The original intent of our study design was to validate the UKB disease signatures in the Copenhagen Hospital Biobank (CHB)(24). We therefore filtered the UKB dataset to only include SNPs that were also present in CHB. Unfortunately, this resource was ultimately not available to us for this validation study, so instead we used the All of Us (AoU) resource, with the limitations that this imposed on the diminished overlap of SNP genotypes.

We also removed SNPs in the low-recombination MHC region of chromosome 6 and conducted LD pruning in PLINK 1.9 (--indep-pairwise 50 2 0.2). This step maximizes opportunities for discovering unique signal by reducing the risk of the combinatorial analytics algorithm redundantly testing effectively equivalent disease signatures comprised of SNPs in linkage disequilibrium. Any meta-GWAS SNPs removed during pruning were added back to the dataset.

The above steps resulted in a UKB cohort with 4,493 cases, 62,574 controls, and 204,423 SNPs used for disease signature discovery.

The distribution of ages differs considerably between cases and controls, with a stronger skew towards older age UKB participants in controls (Supplementary Figure 1). This finding implies that older UKB participants were less likely to have received an ICD-10 code for endometriosis and that a subset of the ‘controls’ in our dataset are likely misphenotyped and may represent missed diagnoses due to historic clinical practices. If that is the case, it would result in decreased associations between genetic variants and a diminished ability to identify biologically relevant disease signatures, as well as reducing the reproducibility rate of the results between datasets (see Limitations of the Analysis).

### Statistical / Conbinatorial Analysis

We ran a combinatorial analysis to identify disease signatures that are significantly enriched in endometriosis cases relative to controls in the UKB discovery cohort. The general methodology behind the hypothesis-free combinatorial analytics approach is described in more detail elsewhere (18,20).

Briefly, the PrecisionLife platform uses a fully deterministic approach to construct and rank disease signatures in ‘layers’ of increasing combinatorial complexity (i.e., adding new SNP-genotypes to the top-ranked disease signatures identified in previous layer). Statistical validation is performed by comparing the properties of disease signatures and their associated ‘networks’ identified by the analysis to the properties of signatures and networks identified in combinatorial analyses using randomly permuted versions of the dataset (i.e., ones constructed by randomly shuffling the case-control assignments to remove any biological link between genotype and phenotype). 1,000 cycles of fully random permutations are typically used.

### Generation of the All of Us Endonetriosis Cohort

We identified a second cohort of female patients diagnosed with endometriosis, along with female controls matched for self-identified ethnicity/race in the AoU Curated Data Repository (CDR) version 8 (accessed on the 29th of January 2025) (25). AoU CDR v8 contains data for over 865,000 American participants including 414,840 patients with whole genome sequencing (WGS) data and 354,400 patients with electronic health record data. Notably, it encompasses a diverse population with a wide age range of 18 to 90 years and substantial representation of non-European ancestry groups, which are frequently underrepresented in genomic research.

Endometriosis cases were identified using the AoU Cohort Browser by selecting all females with whole genome sequencing (WGS) data who had a diagnosis of endometriosis based on ICD9 codes 617.1-9 or ICD-10 codes N80.1-9 and N80.A-D (see Supplementary Table 1). These ICD code criteria are referred to as ‘Source Concepts’ in AoU. This resulted in 4,134 endometriosis cases.

The AoU control cohort was generated by selecting females with WGS data who do not have any evidence of endometriosis, either based on ICD-9/ICD-10 codes (AoU Source Concepts) or SNOMED codes (AoU Standard Concepts) (see Supplementary Tables 2 and 3). We also excluded individuals who have a history of procedures such as laparoscopy or any symptomatic phenotypes potentially consistent with undiagnosed endometriosis, such as pelvic pain, dysmenorrhea, or infertility reported in EHR or surveys (see Supplementary Tables 2 and 3). We used the sex-imputation functionality of PLINK to confirm that all selected case and controls were imputed as genetically female. Applying these criteria, our maximum control population included 191,331 individuals. As in UKB, we observed a similar skew towards higher age among controls than cases in AoU (Supplementary Figure 2).

For each patient, we took whole genome sequence (WGS) data from AoU and extracted all available SNP genotypes included in the filtered UKB genotype dataset. This resulted in 200,578 SNPs available for signature lookups, including 35 of the 42 2023 meta-GWAS SNPs. This lower SNP genotype coverage further restricts the number of matching reproducible signatures that could be found, and this led to 50 signatures being lost from the reproducibility test.

To reduce potential confounding effects arising from population substructure, we matched controls to the endometriosis cases based on self-reported race/ethnicity to generate a dataset with a case:control ratio of 1:4. We employed the probabilistic stratified sampling approach described in (21) to select the subset of 16,536 controls that most closely matches the distribution of demographic subgroups in cases. The demographic breakdown of the AoU endometriosis dataset is included in Supplementary Table 4.

We used principal component analysis (PCA) to model any remaining population substructure within the AoU study cohort. Prior to performing PCA, we first removed all SNPs that are associated with the sex chromosomes, that fall within the MHC region on chromosome 6, or that have minor allele frequency less than 5%. We then conducted LD-pruning in PLINK 1.9 (--indep-pairwise 50 5 0.2) before generating genetic principal components (PCs) using the –-pca command in PLINK 1.9. We selected the top 4 PCs for use in our analyses based on the variance explained by the associated eigenvalues (Supplementary Table 5).

### Statistical Reproducibility of Disease Associations for Combinatorial Signatures in All of Us

We evaluated the degree to which the endometriosis disease signatures identified in the UKB cohort reproduce in AoU, using an approach similar to the one previously used to validate combinatorial disease signatures for long COVID (21). 50 of the disease signatures identified in UKB could not be evaluated in AoU because one or more of their component SNP genotypes were not included in the latter dataset following QC. These were excluded from the analysis.

First, the association between each disease signature and disease status in AoU was assessed via a logistic regression with absence/presence of the disease signature as the dependent variable alongside the top 4 genetic PCs as covariates. The genetic PCs were included as covariates to control for the effects of population substructure on signature frequency.

Using the results of these logistic regressions, we first tested whether any individual signatures are significantly associated with increased risk of endometriosis in AoU, after applying Bonferroni (26) and Benjamini-Hochberg (27) false discovery rate (FDR) adjustments to account for testing of multiple signatures. We then repeated this analysis individually for the output of each ‘layer’ of the UKB combinatorial analysis, where ‘layer’ denotes the number of SNP-genotypes in a signature. We also assessed whether any of the ‘lead’ SNPs identified by the 2023 meta-GWAS (11), which did not include AoU, are significantly associated with endometriosis in our AoU cohort.

Statistically validating replication of disease associations for individual signatures is challenging due to the limited statistical power associated with the relatively small size of the AoU validation cohort. Due to their nature, multi-SNP disease signatures, especially with 4 or 5 SNPs, have much lower frequency than their individual component SNPs, requiring larger sized cohorts to achieve equivalent statistical power. This difference in statistical power is further magnified when the number of disease signatures from combinatorial analysis is orders of magnitude larger than the number of significant loci detected in the meta-GWAS, as is expected when the disease biology reflects a complex network of interacting genes. The greater number of features tested necessitates a much more severe FDR adjustment when assessing statistical significance of the former relative to the latter.

Therefore, we also assessed the evidence for broad reproducibility of positive disease associations among the set of endometriosis signatures in AoU. This enrichment analysis tests whether the proportion of disease signatures that are positively correlated with endometriosis in AoU is significantly greater than expected for a set of signatures unlinked to disease. Observing an enrichment of reproducible signatures would suggest that the set of signatures are reflective of disease biology, even if most individual signatures cannot achieve a level of statistical validation due to the small sample size.

First, we counted the fraction of signatures that have a positive coefficient (odds ratio > 1) in the logistic regression. We call this the ‘reproducibility rate’ for a set of signatures. To test whether the observed enrichment is statistically significant, we randomly reassigned case-control status to the patient cohort while maintaining a fixed case-control ratio, generated logistic regression results in the randomized cohort, and calculated the observed reproducibility rate (i.e., fraction of signatures with odds ratio > 1) in the randomized cohort. Repeating this process 100 times allowed us to generate a distribution of reproducibility rates under the null hypothesis where disease signatures are unlinked to case-control status.

The *p*-value associated with the observed reproducibility data is the fraction of total permutations in which the observed reproducibility rate is equal to or greater than the observed reproducibility rate in the AoU endometriosis cohort. This permutation-based approach for calculating *p*-values is most appropriate because the signatures are non-independent due to shared component SNP-genotypes and therefore violate the assumptions of independent observations required by standard statistical tests.

Our previous analysis of long COVID (21) showed that the reproducibility rates for combinatorial disease signatures were positively correlated with the frequency of those signatures within the population. This likely reflects the reduced statistical power for rare signatures, which are more likely to randomly occur at higher frequency in controls than cases due to random sampling, even when they are biologically associated with disease. Therefore, we assessed the reproducibility rate for endometriosis disease signatures after applying various thresholds for signature frequency in the AoU cohort.

### Reproducibility of Disease Signatures in Traditionally Underserved Populations

We also tested whether the disease signatures identified in the White British UKB cohort reproduced among AoU participants who have self-reported ‘Black’ or ‘African American’ race as well as patients with self-reported ‘Hispanic’ or ‘Latino/a’ ethnicity. These cohorts are not fully mutually exclusive, as they represent responses to different questions within AoU.

The aim of this analysis was to test whether use of combinatorial signatures in a clinical setting can avoid exacerbating historical health inequalities driven by biases in representation, not to identify biological differences between human populations. Therefore, we relied on self-reported race/ethnicity which reflects social constructs, rather than genetic ancestry.

We again included genetic PCs as covariates in the reproducibility analyses for each sub-cohort to control for indirect relationships between signature frequency and disease prevalence resulting from population substructure that could potentially manifest as false positives. We calculated separate PCAs for each sub-cohort using the approach described above for the whole cohort. We then selected the first two PCs as covariates for each ancestry-specific analysis based on the variance explained by the PCA eigenvalues (see Supplementary Table 5).

### SNP and Gene Annotation

SNPs identified in disease signatures were mapped to genes using an annotation cascade process against the human reference genome (GRCh38). SNPs that lie within coding regions of gene(s) were assigned directly to the corresponding gene(s). Remaining SNPs that lie within 2 kb upstream or 0.5 kb downstream of any gene(s) were mapped to the closest gene(s) within this region (28). Additional gene assignments for identified SNPs were made using publicly available cis– or trans-eQTL data for ovary and uterine tissues (29,30) with false discovery rate (FDR) < 0.05.

SNPs located outside the upstream and downstream distance thresholds (2 kb and 0.5 kb respectively) of any gene that did not have any data in relevant tissues were not associated with any gene for biological interpretation. This conservative SNP mapping approach is designed to identify the most likely biologically relevant gene(s) for novel target discovery and repurposing analyses. The 2023 meta-GWAS SNPs that were assigned to a gene in the meta-GWAS manuscript (11) but were not mapped to any genes using this approach are listed in Supplementary Table 6.

The mapped genes were annotated using data from over 50 public data sources (see Supplementary Table 7), and their biological relevance analyzed to develop a deeper understanding of the gene and its potential mechanism of action link to endometriosis phenotypes. All genes were screened for matches against existing drugs in preclinical and clinical development using GlobalData (2024). This was used to identify, evaluate and prioritize existing endometriosis drugs and potential drug repurposing candidates that can be mapped to specific mechanistic patient subgroups identified in this analysis.

### Pathway Enrichnent

Pathway enrichment analysis was performed using the *enrich* function implemented in)the *gseapy* package (32) which tests whether query gene sets are significantly over-represented in the input gene list relative to a background gene universe using a hypergeometric test. Enrichment was assessed across three biological process and pathway databases: Gene Ontology (GO)(33), Reactome (34), and WikiPathways (35). Biological process or pathway *p*-values were adjusted for multiple testing using false discovery rate correction and terms with an adjusted *p*-value lower than 0.05 were reported as significant.

### Reproducibility of Meta-GWAS Disease Associations in All of Us

For comparison, we assessed the replicability and reproducibility of the 35 meta-GWAS SNPs reported in the 2023 study that were included in our AoU dataset. We used the –-logistic command in PLINK 1.9 to generate GWAS results for the AoU cohort, including as covariates the same top 4 genetic PCs used in the reproducibility study for the combinatorial signatures. Although the GWAS implementation in PLINK is not state-of-the-art, it offers an approximate estimate of the degree of reproducible signal from the 2023 meta-GWAS SNPs. We were unable to properly evaluate the replicability and reproducibility of the novel SNPs from the 2026 meta-GWAS as those results were derived in part from AoU.

A disease association for a GWAS SNP is considered to replicate if its logistic regression additive model *p*-value is statistically significant (*p* < 0.05 after Bonferroni FDR adjustment for multiple tests). A disease association for a GWAS SNP is considered to reproduce if the coefficient from the logistic regression additive model is greater than 0 (i.e., odds ratio > 1).

### Phenotype Analysis of Genes using All of Us data

We investigated whether patient subgroups defined by genes mapped to reproduced disease signatures (>1% frequency) were associated with any infertility-related phenotypic features. Four infertility-related phenotypes were defined within the AoU endometriosis cohort for this analysis: (1) female infertility based on in AoU Standard Concept (SNOMEDCode: 6738008), (2) female infertility based on AoU Source Concept using ICD10 code N97, (3) use of any assistive reproductive technology (based on CPT codes 58970, 58974, 58976 and 89250-89259) and (4) a composite phenotype indicating evidence of female infertility based on any of the preceding three criteria. After extraction of these four phenotypes, they were converted into binary features indicating presence (1) or absence (0) of each infertility phenotype.

We performed additional phenotype enrichment analyses by comparing phenotype distributions of patient subgroups linked to a gene to those in the remaining case cohort using Fisher’s exact tests. Multiple testing correction was applied using the Benjamini–Hochberg method to control the false discovery rate (FDR).

## Results

### Results of Conbinatorial Analysis of the UKB Cohort

Combinatorial analysis of the UKB population identified 1,759 multi-SNP signatures significantly associated with endometriosis (UKB disease signatures) containing 3,039 unique SNPs. 1,709 of these signatures could also be assessed in AoU and were retained for the analysis. These signatures include:

- 196 combinations of two SNP-genotypes
- 403 combinations of three SNP-genotypes
- 440 combinations of four SNP-genotypes
- 670 combinations of five SNP-genotypes

Together, these 1,709 combinations include a total of 2,946 unique SNPs, which mapped to 1,309 genes.

Pathways enriched in the disease signatures included cell adhesion, proliferation and migration (36), epithelial-mesenchymal transition (36,37), cytoskeleton remodeling (38), angiogenesis (39,40), as well as signaling pathways such as TGF-β, PI3K/AKT and MAPK (41), many of which have been extensively linked to endometriosis (Supplementary Figure 4).

We then evaluated the frequency and disease association of these UKB disease signatures in AoU. The most common UKB disease signature occurs in 20% of AoU participants, and 20% of UKB disease signatures (342 / 1709) occur in fewer than 1% of AoU participants. Figure 1 presents a histogram of UKB disease signatures by frequency.

**Figure 1.**
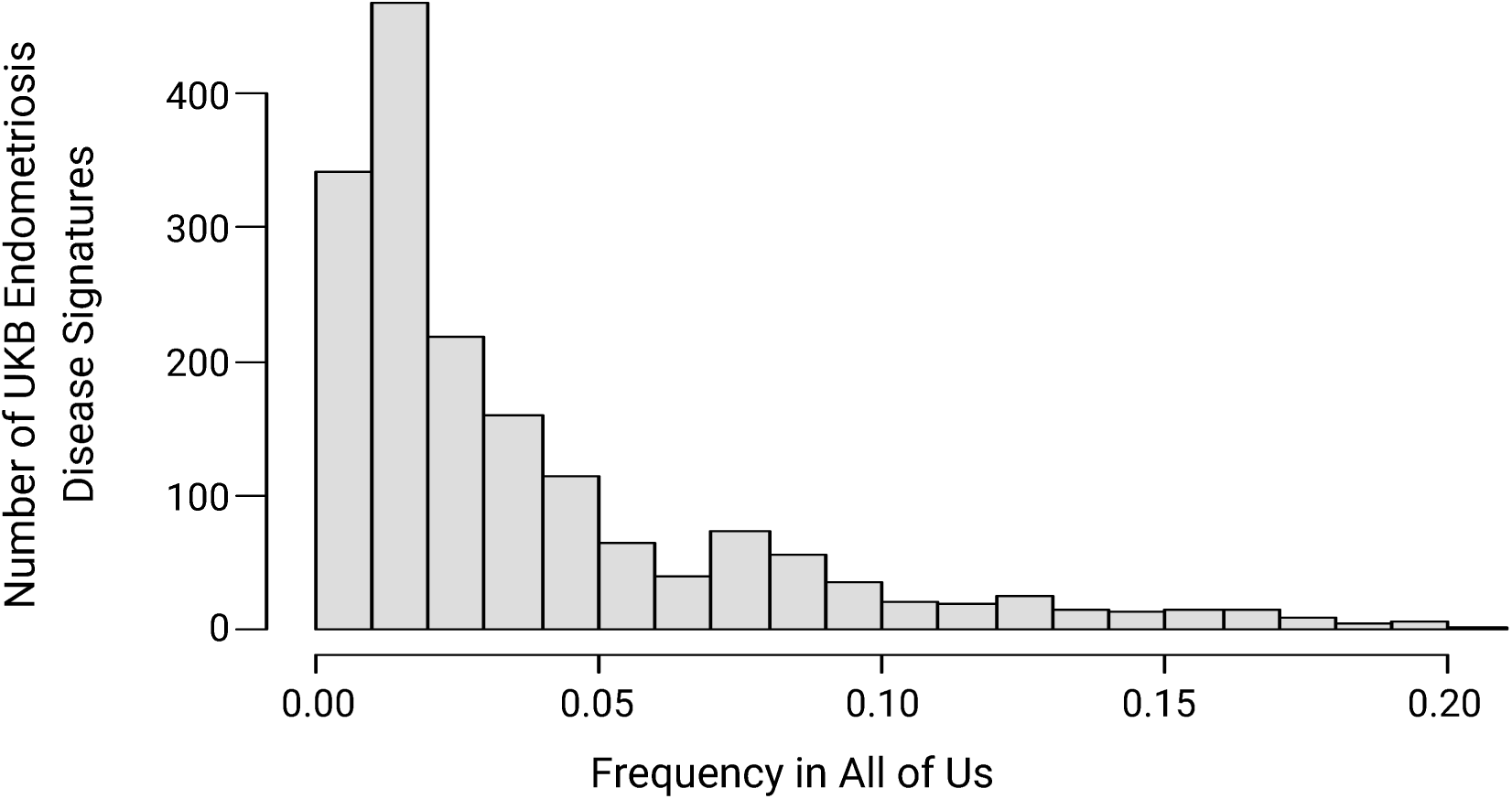
Distribution of UKB endometriosis disease signatures by frequency in AoU.

### Statistical Validation of Individual Disease Signature in All of Us

When considering the full set of 1,709 disease signatures, none were significantly associated with increased disease risk in AoU after applying Bonferroni or Benjamini-Hochberg FDR adjustments. However, when we tested just the 196 disease signatures comprised of 2 SNP-genotypes, resulting in a less severe FDR correction, one signature was significantly associated with increased disease risk in AoU under both FDR adjustment approaches.

This signature, which has a Benjami-Hochberg adjusted *p*-value = 0.038 and logistic regression odds ratio = 1.21 in AoU, is comprised of the heterozygous genotypes for rs11751190 (G/A) and rs1888328 (T/G). The former SNP is located within an intron of the gene *SYNE1* while the latter SNP is located in intronic regions of multiple lncRNAs. *SYNE1* was also linked to a meta-GWAS SNP, but the *SYNE1* SNP in this signature is not the same as the lead *SYNE1* SNP in the meta-GWAS (rs71575922). No disease signatures were statistically significant when we individually assessed 3-, 4-, or 5-SNP genotype signatures.

### Reproducibility of Endonetriosis Disease Associations in AoU Cohort

Evaluating all 1,709 endometriosis disease signatures, we observed a significant enrichment of signatures that also have odds ratios > 1 in AoU (58.5%). This reproducibility rate trends higher when the signatures are filtered by increasingly greater frequency (Figure 2). For example, reproducibility rates increased to 62.8% when assessing the 1,367 signatures with frequency > 1%. The reproducibility rate further increased to 68.1% for the 521 signatures with frequency > 4%, 80.1% for the 176 signatures with frequency > 9%, and 88.3% for the 120 signatures with frequency > 11%. These frequency cutoffs were chosen based on inflection points in reproducibility rates across 1% frequency intervals (e.g., signatures with frequency between 8%-9% vs. 9%-10%). We confirmed that all results are statistically significant, i.e., similar reproducibility rates are observed in fewer than 5% of random permutations (*p* < 0.05, Table 1).

**Figure 2.**
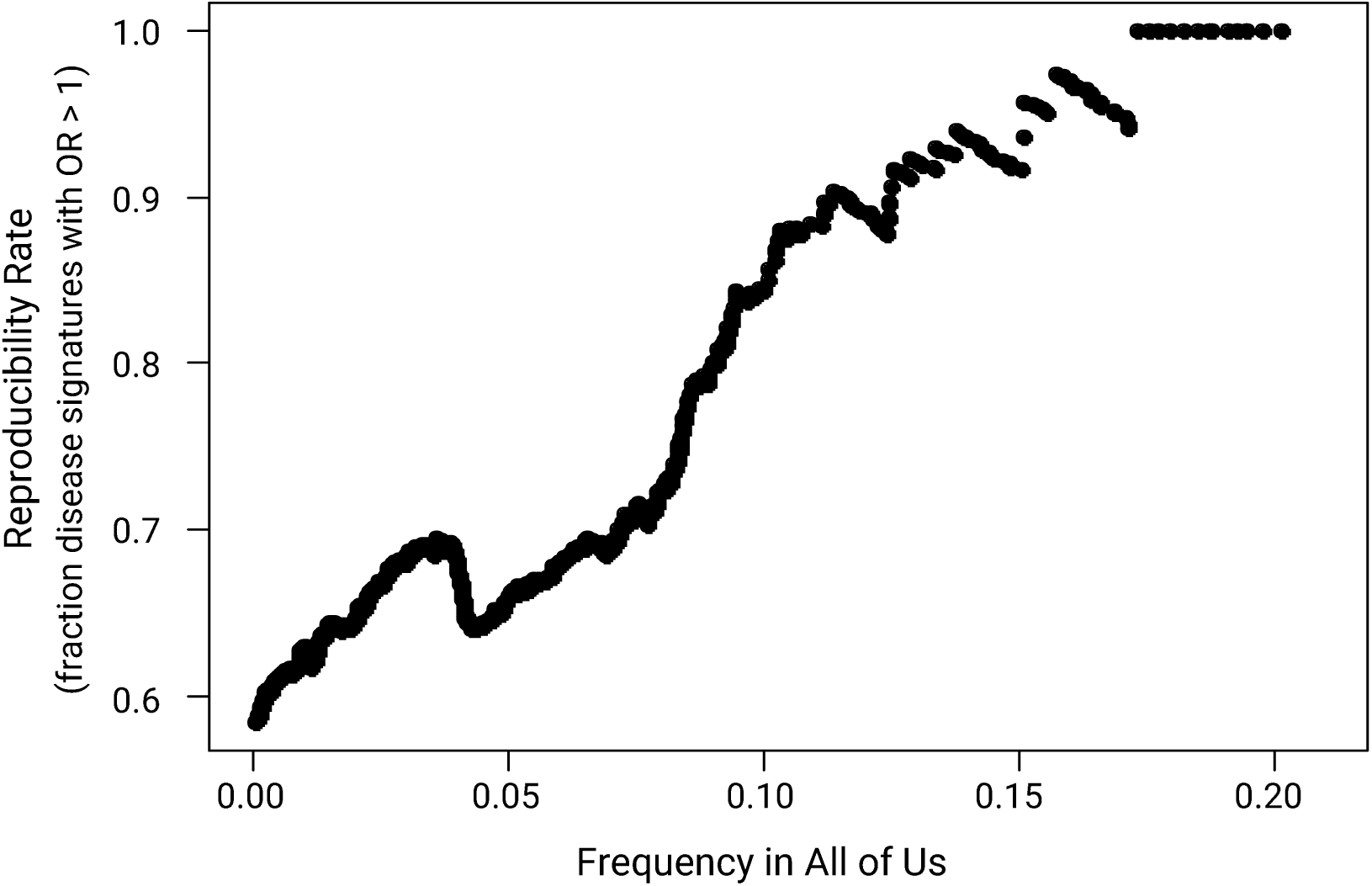
Reproducibility rates for UKB endometriosis disease signatures in AoU using different minimum frequency thresholds.

**Table 1.** Reproducibility statistics in AoU for UKB endometriosis disease signatures using different signature frequency thresholds. Bold indicates statistical significance, *p*-values reported as < 0.01 means that the observed reproducibility was reported in 0/100 random permutations.

| Minimum signature frequency | # signatures tested | % (#) signatures with odds ratio $> 1$ in AoU | % signatures with odds ratio $> 1$ in random permutations mean (range) | $p$-value |
| --- | --- | --- | --- | --- |
| 11% | 120 | 88.3% (106) | 49.5% (14.9% - 83.5%) | **$<0.01$** |
| 9% | 176 | 80.1% (141) | 49.3% (15.2% - 78%) | **$<0.01$** |
| 4% | 521 | 68.1% (355) | 49.7% (24.7% - 70.7%) | **0.04** |
| 1% | 1,367 | 62.8% (859) | 50.4% (33.4% - 64.6%) | **0.02** |
| none | 1,709 | 58.5% (999) | 50.2% (39.4% - 62%) | **0.04** |

UKB disease signatures that reproduce in AoU exhibit a range of odds ratios. Including all reproducing signatures regardless of frequency, 46% have odds ratios greater than 1.1 in AoU, 6% have odds ratios greater than 1.3, the maximum odds ratio is 2.37, and the mean odds ratio is 1.12 (Table 2 and Supplementary Figure 3).

**Table 2.** Odds ratio (OR) statistics for different reproducing UKB endometriosis signature sets and meta-GWAS SNPs in AoU.

| Minimum signature frequency | # reproducing signatures | OR > 1.1 # (%) | OR > 1.3 # (%) | Mean OR | Max OR |
| --- | --- | --- | --- | --- | --- |
| 11% | 106 | 43 (41%) | 0 (0%) | 1.08 | 1.21 |
| 9% | 141 | 57 (40%) | 0 (0%) | 1.08 | 1.21 |
| 4% | 355 | 130 (37%) | 0 (0%) | 1.08 | 1.30 |
| 1% | 859 | 345 (40%) | 11 (1%) | 1.09 | 1.49 |
| none | 999 | 458 (46%) | 60 (6%) | 1.12 | 2.37 |
| meta-GWAS SNPs | 33 SNPs | 5 (15%) | 0 (0%) | 1.06 | 1.29 |

We observed a narrower range of odds ratios and lower mean odds ratios after applying frequency filters, as expected given the trade-off between effect size and feature frequency inherent to disease genetics (Supplementary Figure 3). However, we continue to observe a relatively large proportion of reproducing signatures with moderate odds ratios (> 1.1).

Among reproducing signatures that occur in more than 1% of individuals, the maximum odds ratio is 1.49, the mean odds ratio is 1.09, and 40% of signatures have odds ratio greater 1.1. Among reproducing signatures that occur in more than 9% of individuals, the maximum odds ratio is 1.21, the mean odds ratio is 1.08, and 40% of signatures have odds ratios greater than 1.1. In comparison, the mean odds ratio for the set of 2023 meta-GWAS SNPs that reproduce in AoU (33 of 35) is 1.06 and just 15% have odds ratios greater than 1.1.

### Reproducibility Across Self-Identified Race/Ethnicity Cohorts

Reproducibility rates for all combinatorial UKB disease signatures in self-identified ‘Black / African American’ and ‘Hispanic / Latino’ cohorts from AoU are similar to reproducibility rates for the whole cohort (Table 3). We also observe similar reproducibility rates across cohorts for signatures with greater than 1% frequency and greater than 4% frequency. Reproducibility rates for signatures with greater than 9% frequency are lower than the reproducibility rates for the full cohort but still demonstrate strong enrichment of reproducing signatures.

**Table 3.**
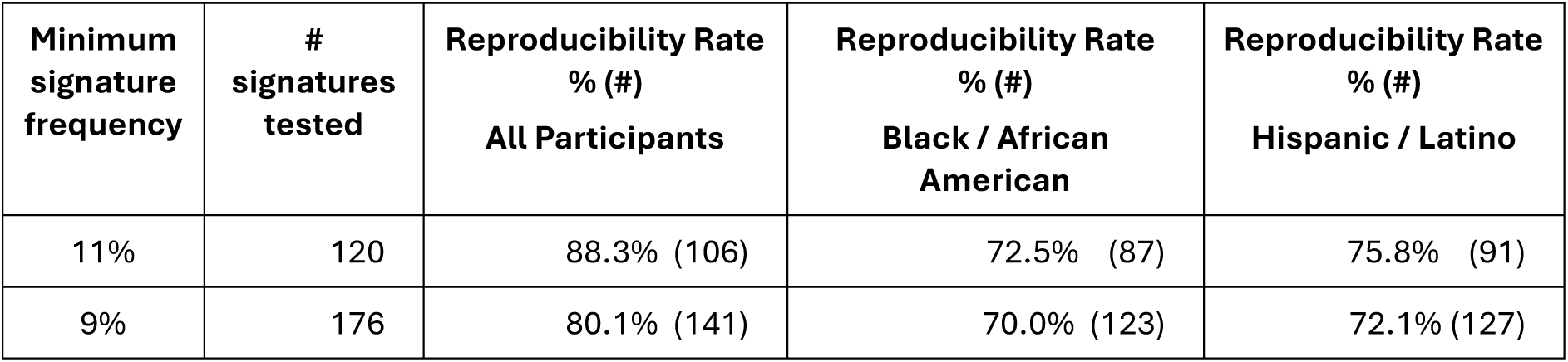
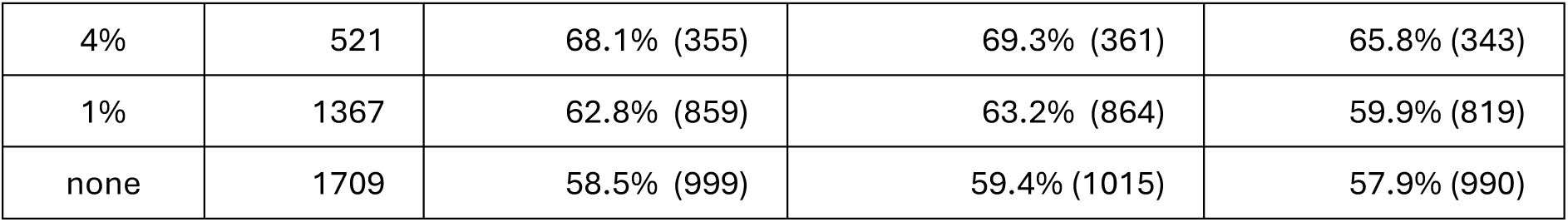
Reproducibility statistics under different signature frequency thresholds for UKB endometriosis disease signatures in self-identified ‘Black / African American’ and ‘Hispanic / Latino’ AoU cohorts compared to all participants.

### Replicability and Reproducibility of Meta-GWAS Results

35 of the 42 meta-GWAS SNPs reported in the 2023 study were present in the AoU dataset used in our replication/reproducibility analysis. The identified risk alleles for 6 (17%) of these meta-GWAS SNPs, annotated to the genes *CDKN2-BAS1*, *FRMD7*, *LINC00629*, *WNT4*, *MLLT10*, and *KDR*, are also significantly associated with increased risk of endometriosis in AoU based on GWAS results after applying a Bonferroni FDR correction (see Supplementary Table 8). An additional 4 SNPs, annotated to the genes *CD109*, *ACTL9*, *GDAP1*, and *VEZT* significantly replicate in AoU if we apply a Benjamini-Hochberg FDR correction, corresponding to a cumulative 29% replication.

The GWAS associations for 21 (60%) of the 35 meta-GWAS SNPs are not even nominally significant in AoU (*p* > 0.05), including 9 SNPs (26%) with *p*-values > 0.5. This lack of statistical replication likely reflects the much smaller sample size of the cohorts for AoU vs. the meta-GWAS, as 94% (33/35) of the meta-GWAS SNPs reproduce with odds ratios greater than 1.

20% of the UKB disease signatures assessed in AoU (336 / 1709) contain at least one of the 2023 meta-GWAS SNPs. 17 of the 35 meta-GWAS SNPs are present in at least one disease signature (Table 4). Disease signatures assigned to 11 of these 17 meta-GWAS SNPs have reproducibility rates greater than 60% in the full AoU cohort, and the overall reproducibility rate for these 336 signatures is 74% (247 / 336).

**Table 4.** Reproducibility of UKB disease signatures linked to meta-GWAS SNPs and their associated genes in the 2023 meta-GWAS study.

| Meta-GWAS gene(s) | Lead 2023 meta-GWAS SNP | # signatures with lead meta-GWAS SNP | Reproducibility rate of meta-GWAS SNP signatures in AoU (OR > 1) | # Signatures assigned to meta-GWAS gene | Reproducibility rate of meta-GWAS gene signatures in AoU (OR > 1) |
| --- | --- | --- | --- | --- | --- |
| *ABO* | rs507666 | 2 | 100% | 2 | 100% |
| *ACTL9* | rs2967684 | 2 | 100% | 2 | 100% |
| *ASTN2* | rs10983311 | 0 | n/a | 3 | 67% |
| *CD109* | rs4540228 | 4 | 100% | 4 | 100% |
| *CDKN2B-AS1* | rs10122243 | 4 | 25% | 30 | 53% |
| *CEP112* | rs7214750 | 0 | n/a | 1 | 100% |
| *DLEU1* | rs7334326 | 1 | 100% | 3 | 67% |
| *DNM3* | rs2421985 | 0 | n/a | 3 | 100% |
| *FRMD7* | rs5933091 | 2 | 100% | 2 | 100% |
| *GDAP1* | rs10090060 | 1 | 100% | 2 | 100% |
| *GREB1* | rs11674184 | 3 | 33% | 19 | 53% |
| *IGF1* | rs10860864 | 3 | 33% | 3 | 33% |
| *FSHB/ARL14EP* | rs3858429 | 34 | 56% | 35 | 57% |
| *KDR* | rs1903068 | 73 | 45% | 73 | 45% |
| *MLLT10* | rs10828249 | 2 | 50% | 2 | 50% |
| *RIN3* | rs57281976 | 0 | n/a | 2 | 50% |
| *RNLS* | rs7907732 | 1 | 100% | 1 | 100% |
| *SKAP1* | rs66683298 | 14 | 64% | 15 | 67% |
| *SYNE1* | rs71575922 | 34 | 74% | 295 | 80% |
| *WNT4/CDC42* | rs10917151 | 156 | 92% | 156 | 92% |
| 7p12.3 | rs55909142 | 9 | 67% | 10 | 60% |

If we expand the definition of meta-GWAS signatures to include any signature containing a SNP that is annotated to one of the meta-GWAS genes, the number of 2023 meta-GWAS signatures increases to 644 (38% of signatures) with a reproducibility rate of 75%. The number of associated 2023 meta-GWAS genes increases by 4 to a total of 21, including two genes where the meta-GWAS SNP was not detected in the combinatorial analysis and two where the meta-GWAS SNP was absent from the AoU dataset.

The increase in meta-GWAS signatures when considering alternative SNPs is largely due to the inclusion of 261 signatures that contain one of three SNPs assigned to *SYNE1* that differ from the lead meta-GWAS SNP. The 80% reproducibility rate for the 261 signatures associated with these alternative *SYNE1* SNPs is higher than the 74% reproducibility rate for signatures associated with the lead *SYNE1* meta-GWAS SNP.

Of the 71 novel lead SNPs from the 2026 meta-GWAS, 10 were present in the UKB dataset used for the combinatorial analysis. None of these novel meta-GWAS SNPs occur in the set of disease signatures from our study. However, 9 novel genes mapping to 2026 but not 2023 meta-GWAS SNPs map to the same genes as SNPs in the disease signatures,

Reproducibility rates in AoU for the 36 signatures mapping to these novel meta-GWAS gene are generally low (53%, Supplementary Table 9) in comparison to the 2023 meta-GWAS signatures, even though the 2026 meta-GWAS was derived in part from AoU samples.

667 signatures (39% of 1709 signatures) contain at least one SNP mapping to the same gene as a 2023 or 2026 meta-GWAS SNP. Together these meta-GWAS signatures have a reproducibility rate of 74%.

### Reproducible Novel Gene Findings

The combinatorial analysis identified novel associations between SNPs and endometriosis in two contexts: novel SNPs that occur in the same disease signature as at least one meta-GWAS SNP (‘meta-GWAS signatures’) and novel SNPs that occur in disease signatures containing no meta-GWAS SNPs (‘non-meta-GWAS signatures’). SNPs can also occur in a mix of meta-GWAS and non-meta-GWAS signatures.

Higher frequency disease signatures exhibited the strongest evidence for a consistent link to endometriosis in the AoU replication analysis. A total of 151 unique SNPs mapping to 99 genes (see Extended Tables 1 and 2) were identified among the 141 disease signatures that have frequency > 9% and are reproduced in AoU (odds ratio > 1, p < 0.01). 7 of these 99 genes were previously identified in the 2023 meta-GWAS. 13 additional genes have been previously associated with endometriosis according to Open Targets (42) or Pharos (43), providing independent confirmation of a potential role in disease, but were not identified by the meta-GWAS (see Supplementary Table 10). The remaining 79 candidate genes represent novel disease-associated genes. 33 of these are tractable drug targets (see Extended Table 3).

Of the 99 candidate genes mapping to high frequency disease signatures, more than 50% are expressed in endometrium and ovary (nTPM > 1; see Extended Table 4). Pathways enriched in these candidate genes included cell-adhesion and proliferation mechanisms, angiogenesis along with neurological processes (Supplementary Figure 5, Extended Table 5). Many of these novel genes occur in meta-GWAS signatures which exhibit strong reproducibility rates overall, but it is challenging to quantify the contribution of meta-GWAS genes vs. novel genes to this reproducibility.

To validate the ability of combinatorial analytics to identify novel reproducible candidate genes associated with endometriosis, we focused on the set of 25 SNPs that were found in 25 or more disease signatures (termed ‘top SNPs’). This criterion reflects the network geometry-based approach used in the PrecisionLife combinatorial analytics platform, which more strongly supports SNPs that are repeatedly linked to disease status in different combinatorial contexts.

20 of these 25 top SNPs were annotated to genes. 6 of those 20 genes overlap with genes identified by the 2023 meta-GWAS (the meta-GWAS SNPs annotated to *WNT4*/*CDC42*, *CDKN2B-AS1*, and *FSHB/ARL14EP*, plus 3 SNPs associated with *SYNE1* including the lead meta-GWAS SNP). Reproducibility statistics for these candidate genes are included in Table 4 and Supplementary Table 8). None of these 20 genes overlap with the novel genes identified by the 2026 meta-GWAS.

Of the remaining 14 annotated top SNPs, 9 most frequently occur (> 50%) in non-meta-GWAS disease signatures (i.e., signatures that contain no component SNPs annotated to a meta-GWAS gene). We refer to these 9 SNPs as ‘top novel SNPs’. Importantly, all 9 top novel SNPs exhibit a strong enrichment of disease signatures that reproduce in AoU (Table 5). Reproducibility rates for the top novel SNPs are especially strong for non-meta-GWAS signatures, ranging between 72% to 85%. Reproducibility of these non-meta-GWAS signatures unambiguously demonstrates the capability of combinatorial analytics to identify novel signal missed by GWAS.

**Table 5.**
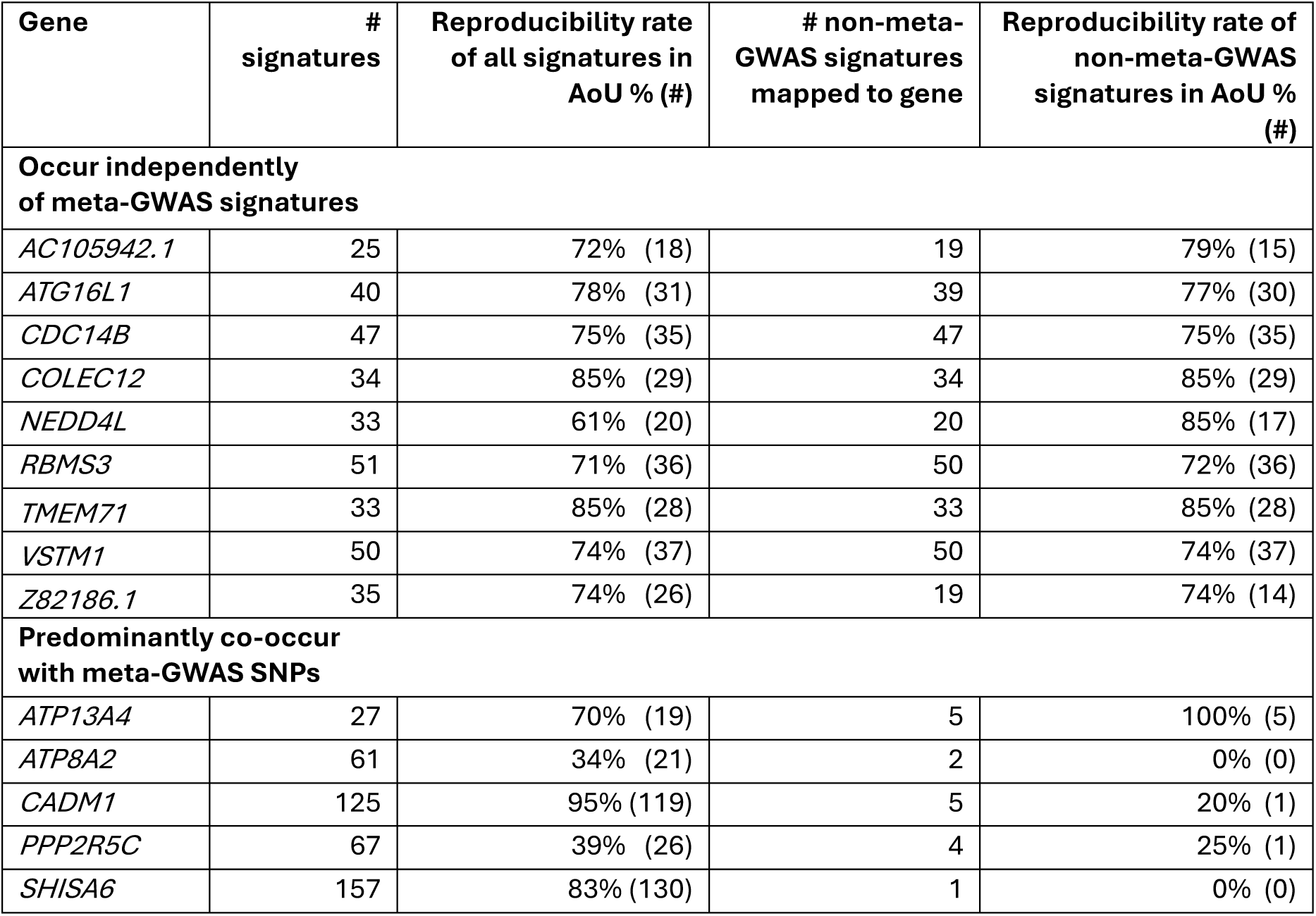
Reproducibility rates in AoU endometriosis cohort for gene-annotated non-meta-GWAS SNPs that occur in 25 or more disease signatures (‘top SNPs’).

| Gene | # signatures | Reproducibility rate of all signatures in AoU % (#) | # non-meta-GWAS signatures mapped to gene | Reproducibility rate of non-meta-GWAS signatures in AoU % (#) |
| --- | --- | --- | --- | --- |
| **Occur independently of meta-GWAS signatures** |  |  |  |  |
| *AC105942.1* | 25 | 72% (18) | 19 | 79% (15) |
| *ATG16L1* | 40 | 78% (31) | 39 | 77% (30) |
| *CDC14B* | 47 | 75% (35) | 47 | 75% (35) |
| *COLEC12* | 34 | 85% (29) | 34 | 85% (29) |
| *NEDD4L* | 33 | 61% (20) | 20 | 85% (17) |
| *RBMS3* | 51 | 71% (36) | 50 | 72% (36) |
| *TMEM71* | 33 | 85% (28) | 33 | 85% (28) |
| *VSTM1* | 50 | 74% (37) | 50 | 74% (37) |
| *Z82186.1* | 35 | 74% (26) | 19 | 74% (14) |
| **Predominantly co-occur with meta-GWAS SNPs** |  |  |  |  |
| *ATP13A4* | 27 | 70% (19) | 5 | 100% (5) |
| *ATP8A2* | 61 | 34% (21) | 2 | 0% (0) |
| *CADM1* | 125 | 95% (119) | 5 | 20% (1) |
| *PPP2R5C* | 67 | 39% (26) | 4 | 25% (1) |
| *SHISA6* | 157 | 83% (130) | 1 | 0% (0) |

The other 5 top SNPs predominantly co-occur with meta-GWAS SNPs (in > 80% of signatures), and as such we cannot evaluate whether reproducibility or lack thereof is due to the core SNP or the meta-GWAS SNP. Of these meta-GWAS signatures, 3 show high rates of reproducibility, while 2 show no evidence of reproducibility (Table 5).

### Association of Genes with Infertility Phenotypes

We investigated whether specific patient subgroups associated with genes that reproduced in AoU were associated with infertility phenotype in the dataset. The analysis highlighted 37 genes that showed the strongest associations across three AoU infertility phenotypes (see Supplementary Table 11, Supplementary Figure 6), however, due to the high multiple-testing burden and limited phenotype data (13.47%, Supplementary Table 12), no association passed the FDR threshold. These gene-infertility associations are reported only for exploratory and hypothesis-generating purposes.

Many infertility-associated genes identified in this analysis appear to be novel in the context of both endometriosis and female infertility, necessitating further functional investigation to elucidate the underlying biological mechanisms. Among genes with prior support in the literature, several are implicated in female reproductive hormone signaling pathways (e.g. *LRP1B* (44), *ETV1* (45)) and in processes related to endometriotic lesion development and proliferation (e.g. e.g. *DEC1* (*46*)*, CCDC3* (47)).

## Discussion

Combinatorial analysis identified 1,709 combinatorial disease signatures that are associated with significantly increased prevalence of endometriosis in a UKB White British cohort. These UKB endometriosis disease signatures exhibited a significant enrichment of reproducible disease signal in AoU. That is, we observed significantly more UKB disease signatures that are also positively correlated with increased prevalence of endometriosis in AoU than expected by chance. These results include many new genes and mechanisms, and the broader biological context captured by these combinatorial disease signatures provides potential to improve our understanding of disease biology and build clinical development tools to support novel therapeutics programs.

Reproducibility rates are especially high for higher-frequency signatures, which is encouraging for precision medicine applications as signatures with higher frequency offer potential clinical utility across more patients. The increased reproducibility of higher-frequency signatures is consistent with observations from a reproducibility study of long COVID disease signatures (21) and likely reflects the lower sampling variance associated with increased sample sizes when estimating the signatures’ odds ratios. That is, the observed odds ratio of biologically relevant low-frequency signatures is more likely to be less than 1 due to random sampling effects.

Despite using a much smaller dataset, combinatorial analysis identified many of the same gene-disease associations previously identified by a large scale meta-GWAS (11) as well as several novel genetic associations with endometriosis. The latter include many genes that have previously been linked to endometriosis in non-GWAS studies, providing the first genetic support for their role in disease.

We identified 79 novel gene associations that have high (> 9%) frequency and reproduce in the independent, ancestrally diverse AoU dataset, with 3 candidate genes overlapping and 6 additional candidate genes associated with the highly reproducible top novel SNPs. Although none of the 9 top candidate genes have been previously linked to endometriosis, several play critical roles in pathways and biological processes that have been hypothesized to be important to its disease biology.

### First Genetic Evidence in Endonetriosis for Many Genes

Identification of genes previously linked to endometriosis via non-genetic studies also provides support for the biological validity of the combinatorial analysis results. For example, we identified disease signatures linked to serotransferrin (TF), an iron binding protein which transports Fe^3+^ ions from sites of absorption and heme degradation to where it is utilized or stored.

Dysregulation of iron homeostasis is believed to be involved in endometriosis (48). Elevated levels of iron occur in endometriotic tissues (48–50) and these have been associated with inflammation, oxidative stress, cellular damage, and ferroptosis (51). TF levels are elevated in peritoneal fluid from women with endometriosis (52,53) and TF saturation is correlated with disease (54). Populations of stromal cells and macrophages from endometrial tissues express elevated levels of the transferrin receptor 1, which is responsible for cellular uptake of TF and iron, resulting in increased iron loading (49,55).

Serotransferrin and antioxidants provide protection for mouse oocytes against the oxidative damage and dysmaturity induced by ovarian follicular fluid from endometriosis patients (56,57). Our observations lend support to the hypothesized importance of iron management in endometriosis and its potential utility for therapeutic manipulation.

Additional examples of genes with previous non-genetic associations are listed in Extended Table 2.

### Novel Insights into the Genetics of Endonetriosis

We identified 9 novel candidate endometriosis genes associated with the set of top SNPs that feature most prominently in non-meta GWAS disease signatures, all of which universally exhibit high degrees of reproducibility in AoU. This subset includes multiple candidate genes associated with autophagy and macrophage biology as well as others that could hold significant potential for novel therapeutics and targeted drug repurposing/repositioning.

### Autophagy

Autophagy, a cellular process responsible for degradation and recycling of intracellular components (58), has previously been implicated in the pathophysiology of endometriosis. Autophagic activity is suppressed in ectopic endometrial tissue, facilitating the survival, immune evasion, and proliferation of ectopic cells (59–61). Among the 9 top novel candidate genes identified in our study, *ATG16L1* and *NEDD4L* both play central roles in autophagy.

*ATG16L1* encodes a core autophagy protein that forms a complex with ATG5 and ATG12, acting as an E3-like ligase essential for substrate recognition and autophagosome biogenesis (62). While *ATG16L1* has not previously been associated with endometriosis, knockdown of interacting autophagy genes *ATG5* and *ATG7* has been shown to impair decidualization in human endometrial stromal cells (63), suggesting a potential role in endometrial function and disease.

*NEDD4L* codes for an E3 ubiquitin ligase that regulates autophagy by targeting the proteins ULK1 and ASCT2 for ubiquitination and degradation (64). Moreover, NEDD4L has been reported to inhibit the activity of mTOR, a key negative regulator of autophagy (65). Although *NEDD4L* represents a novel association with endometriosis, a related E3 ligase NEDD4 has been shown to inhibit ferroptosis and promote endometrial lesion stromal cell survival by mediating the ubiquitination and degradation of PTGS2 (66).

Notably, both *ATG16L1* and *NEDD4L* have been implicated in inflammatory bowel disease (IBD) (67,68). The *ATG16L1* SNP identified in this study was also strongly associated with Crohn’s disease in a published GWAS (67). This finding is consistent with the reported symptomatic overlap and high rates of co-prevalence between Crohn’s disease and endometriosis (69). Collectively, this evidence suggests that dysregulation of *ATG16L1* and *NEDD4L* may contribute to impaired autophagic homeostasis in endometriosis, potentially supporting lesion development, inflammation, and disease progression.

### Macrophage Biology

Macrophage dysfunction has been hypothesized to contribute to the development and progression of endometriosis (70). Peritoneal macrophages produce more *VEGF* in endometriosis patients relative to healthy individuals, and transcriptomic analyses highlight macrophage-derived cytokines as key drivers of inflammation in endometriosis (71,72). In mouse models, macrophage depletion reduces lesion size, vascularization, and inflammatory pain, underscoring their central role in disease development (73,74). During menstruation, macrophages clear endometrial debris via scavenger receptor–mediated phagocytosis (75). Intriguingly, one of the top novel candidate genes identified in our analysis, *COLEC12*, encodes one of these scavenger receptors involved in host defense and phagocytosis.

A second top novel candidate gene, *VSTM1*, encodes an immunoglobulin superfamily protein that enhances IL17A secretion by CD4+ T cells (76). Although *VSTM1* itself has not been studied in endometriosis, *IL17A* is known to drive pathogenic M2 macrophage polarization in lesions (77). Both genes show macrophage-enriched expression (78), and while their altered expression in endometriosis remains unconfirmed, dysfunction in either may impair phagocytosis, sustain inflammation, or disrupt tissue remodeling, promoting lesion persistence.

### Novel Drug Discovery and Repurposing Targets

Alongside less well-studied but potentially druggable novel candidate genes, the reproducing disease signatures for endometriosis include several genes whose protein products are targets for well-established therapeutics that have not previously been explored in the treatment of endometriosis, i.e., drug repurposing opportunities. We ranked these drug repurposing candidates based on several criteria, including a mechanistic link to the disease, target selectivity, appropriate directionality of modulation, and a modality and safety profile suitable for endometriosis patients (23).

One example of a high-ranked candidate drug repurposing target included in the endometriosis disease signatures is the peptidase angiotensin-converting enzyme (ACE). ACE plays a major role in the renin-angiotensin-aldosterone system (79), which regulates sodium retention by the kidney and blood pressure by converting the peptide hormone angiotensin I (AT1) to angiotensin II (AT2), resulting in an increase of the vasoconstrictor activity (80). Although *ACE* was not identified in the endometriosis meta-GWAS, the product of ACE activity, AT2, enhances degradation of extracellular matrix proteins in cultures of human endometrial stromal cells, while also improving their viability and increasing proliferation and migration (81). ACE inhibitors (captopril, ramipril) reduce the growth of endometrial implants in rat models (82,83) and the expression of the receptors for AT2 (AGTR1, AGTR2) is altered in endometrial lesions (84,85).

ACE inhibitors have been in widespread clinical use since the 1980s for a variety of indications including coronary artery disease, congestive heart failure, chronic kidney disease and myocardial infarction (86). A link between endometriosis and hypertension has previously been identified (87–89) but the opportunity to exploit existing therapeutic modulators of ACE or the angiotensin II receptor inhibitors has not been investigated.

Many other shortlisted repurposing candidates identified offer clear mechanistic hypotheses for their roles in lesion development, inflammation, fibrosis and – in a few cases – endometriosis-associated infertility. Future work based on these results will prioritize the drug repurposing candidates (see Extended Table 2) mapped to high frequency disease signatures as described above. Candidates will only be selected if they correspond to a clear hypothesis for how they might benefit endometriosis patients safely. Additional research will investigate the potential for using the disease signatures identified in this study as biomarkers to stratify patients by mechanistic subtype and match patients with drugs that target their unique disease mechanisms.

### Infertility Phenotype Association Analysis

The proportion of endometriosis cases within the AoU cohort with at least one additional infertility diagnostic code was only 13.47% (n = 557) (Supplementary Table 11). In contrast, epidemiological studies have estimated that the prevalence of female infertility among women with endometriosis ranges from 25% to 50% (90,91).

This discrepancy suggests that a substantial proportion of endometriosis cases within AoU may have experienced infertility that has not been captured in electronic health records (EHRs), either due to underdiagnosis or incomplete reporting. Consequently, the observed associations between endometriosis-associated genes and infertility phenotypes are likely to be underestimated in the AoU dataset. These findings underscore the need for more comprehensive endometriosis cohorts incorporating detailed, longitudinal phenotypic data capable of capturing clinically meaningful outcomes for affected individuals.

Future work will extend this analytical framework to genomic datasets from other chronic women’s health conditions associated with an increased risk of infertility, such as polyendocrine metabolic ovarian syndrome (PMOS, formerly PCOS). This approach will enable assessment of whether the identified genetic associations are specific to endometriosis-related infertility or reflect broader mechanisms underlying female infertility.

Given that fertility outcomes represent a primary concern for individuals with endometriosis (92), we will further evaluate the predictive performance of the identified disease signatures in larger, independent cohorts, including those enriched for endometriosis-associated infertility. Improved understanding of the genetic architecture of infertility in endometriosis and related conditions may inform more effective patient stratification, earlier diagnosis, and the development of personalized approaches to fertility management.

### Linitations of the Analysis

We observed a significant enrichment of reproducible disease signatures in the output of the combinatorial analysis, illustrating the potential suitability for applying these disease signatures in aggregate to provide insights into the biology of endometriosis. However, it was only possible to statistically validate replication of a single individual disease signature, preventing us from confidently asserting which of the reproducible disease signatures are most important for disease biology.

Statistically validating individual disease signatures in an independent dataset is challenging due to the small size of available cohorts, the large number of signatures, the relative rarity of individual signatures, and the complexity of disease biology. This is illustrated by the fact that fewer than 30% of the meta-GWAS SNPs replicate significantly in AoU, despite higher frequencies, much less biological complexity, and orders of magnitude fewer features and correspondingly reduced need for FDR adjustment to account for multiple tests relative to the replication analysis required for disease signatures. In contrast, 94% of the meta-GWAS SNP risk alleles are also positively correlated with increased prevalence of endometriosis in AoU, suggesting that the lack of broad statistical replication for both sets of results is likely due to lack of statistical power provided by the relatively small cohort.

Reliance on patient data provided by UKB and AoU also introduces several potential limitations for this study. Both datasets have relatively small populations of diagnosed endometriosis cases and have material levels of phenotypic misclassification in both cases and controls as is common in complex diseases. In addition, the UKB genotyping array provides limited SNP coverage for identification of disease signatures. Each of these factors are expected to reduce the number of UKB disease signatures found during the initial combinatorial analysis and the level of their reproducibility reported in AoU.

A key limitation is the necessary reliance on ICD-10 coding, which is often inconsistently and inaccurately applied, to identify cases and controls. Surgical confirmation is needed to reliably identify patients with endometriosis, and such data is not consistently available in UKB or AoU. As a result, some participants with ICD-10 codes for endometriosis may not actually have the disease. The observed negative correlation between age and endometriosis in both datasets is likely to be an artifact of high prevalence of mis-/undiagnosed cases amongst older women due to lack of recognition of the disease and use of older, less reliable diagnostic criteria when they were younger. This pattern suggests that a subset of the ‘controls’ in the UKB and AoU study cohorts likely represent misphenotyped patients.

Phenotypic misclassifications typically reduce observed effect sizes by artificially increasing the resemblance between cases and controls, and this issue is equally problematic for GWAS (93). Such misclassification also poses a challenge for reproducibility analyses, as signal dilution lowers the statistical power of results (94). For example, it raises the likelihood that a biologically meaningful disease signature could display an odds ratio below 1 simply because of random sampling, as shown in (21).

Accordingly, we anticipate that phenotypic misclassification in UKB will have suppressed the number of statistically significant signatures found during the combinatorial analysis. Phenotypic misclassification within AoU will then have suppressed the number of signatures found to reproduce to a statistically significant degree. This means that the reproducibility results reported here likely represent a lower-bound estimate of the reliability of the combinatorial analytics approach.

Although more robustly phenotyped genomic datasets of endometriosis patients are available, they do not include sufficient controls. Many genetic association studies merge case data with data for controls that were genotyped as part of different studies. However, differences in sample collection, genotyping, and SNP calling between studies can introduce technological ‘batch effects’, i.e., significant differences in allele frequencies between cases and controls related to dataset preparation rather than the disease status of the individuals. When this occurs, signatures that are indirectly associated with disease due to batch effects in the data can be very difficult to separate from true biological associations (19,95). Any benefits to this study that could be gained from using better phenotyped case data would therefore likely be outweighed by the reduced confidence in the observed disease associations.

Finally, our phenotypic enrichment analysis was constrained by the limited availability of endometriosis-specific clinical data available for biobank datasets. Endometriosis disease staging data was not available and although infertility phenotype could be partially evaluated, the available data was insufficient for robust inference.

### Wider Inplications of the Findings

The study’s findings include several candidate genes and mechanisms novel to endometriosis. This is important to provide new avenues of research to find new clinical options for a disease that is currently poorly diagnosed and whose main therapies are surgery and hormone treatment, neither of which is particularly effective or appropriate for many women (96).

Many of these genes are associated with targets or mechanisms that are plausible for the disease given evidence from other studies, and which can be modulated using known safe and well-tolerated active compounds. These can therefore be validated relatively readily using existing drug development candidates (repositioning) or on-market/generic compounds (repurposing) in small proof-of-concept human studies. Such studies would ideally be guided by genetic biomarkers used to recruit patients whose disease is driven by that mechanism.

These would provide rapid and cost-effective clinical validation of the disease modifying potential of the novel targets and complementary diagnostic tools to select patients most likely to respond to these drugs in a clinical setting.

Given the high degree of reproducibility, these same genetic/mechanistic biomarkers can also potentially be used during novel drug development to build a precision regulatory strategy, and to recruit likely responders into clinical trials, accelerating and derisking their clinical development.

When fully developed and validated, such detailed genetic insights could also potentially be used in a clinical setting for rapid and non-invasive differential triage of patients presenting with deep pelvic pain to facilitate quicker referral of women to achieve a definitive diagnosis, and to evaluate patient prognosis for severity and progression to infertility, endometriomas, and other complications.

### Supporting Health Equity

It is important that the results of genomic analyses support health equity across as many patients as possible, including those with different ancestries. GWAS results and related risk scores are often derived predominantly from European cohorts and may fail to transfer well across populations (97–100).

Better population coverage in the datasets used for genetic analyses is widely recognized as important for ensuring that study findings and derived tools do not disadvantage populations of patients, but the choice of analytical methods is often overlooked as a critical factor in finding transferable results that reproduce well across patient with different ancestries and other epidemiological features (101).

Although the initial combinatorial analysis was performed on a White British only cohort, the reproducibility results from this study are broadly consistent across all AoU participants regardless of their self-reported race/ethnicity. This observation is crucial as it implies that these disease signatures can be used to inform precision medicine healthcare without adversely affecting historically underserved populations and further increasing health disparities.

## Conclusions

The high level of reproducibility of combinatorial disease signatures across different ancestries, based on combinations of relatively common variants, and the finding of a large number of both novel and known genes is encouraging for furthering the study of endometriosis and its clinical care.

Further combinatorial analysis approaches in endometriosis would benefit from access to larger datasets with even wider population diversity, more secure diagnosis, more harmonized health/symptom surveys, and deeper whole genome, longitudinal clinical, proteomic, and metabolic data. We hope that such datasets will be made available for deeper study in the future.

Much more work is needed in endometriosis; a rapid and accurate test for the disease would enable patients to access timely care and reduce the potential for years of misdiagnosis, suffering and clinical waste, but it also requires there to be a range of good precision therapeutic options once those women are diagnosed. Progress on both fronts will help strategically prioritize much needed women’s health research initiatives to make more rapid progress in addressing this massive global challenge and improving patients’ lives.

## Ethics Approval and Consent to Participate

Research described in this article has been conducted using data from All Of Us Research Program and UK Biobank (application number 44288).

UK Biobank has approval from the North West Multi-centre Research Ethics Committee (MREC) as a Research Tissue Bank (RTB) approval, and researchers do not require separate ethical clearance and can operate under this RTB approval.

Institutional Review Board (IRB) approval was obtained prior to enrollment of patients in the All of Us Research Program. Informed consent for all participants is conducted in person or through an eConsent platform that includes primary consent, HIPAA Authorization for Research use of EHRs and other external health data, and Consent for Return of Genomic Results. The protocol was reviewed by the Institutional Review Board (IRB) of the All of Us Research Program (IRB Approval Date: Dec 03, 2021). The All of Us IRB follows the regulations and guidance of the NIH Office for Human Research Protections for all studies, ensuring that the rights and welfare of research participants are overseen and protected uniformly. The All of Us Research Program is supported by the National Institutes of Health, Office of the Director: Regional Medical Centers (OT2 OD026549; OT2 OD026554; OT2 OD026557; OT2 OD026556; OT2 OD026550; OT2 OD 026552; OT2 OD026553; OT2 OD026548; OT2 OD026551; OT2 OD026555); Inter agency agreement AOD 16037; Federally Qualified Health Centers HHSN 263201600085U; Data and Research Center: U2C OD023196; Genome Centers (OT2 OD002748; OT2 OD002750; OT2 OD002751); Biobank: U24 OD023121; The Participant Center: U24 OD023176; Participant Technology Systems Center: U24 OD023163; Communications and Engagement: OT2 OD023205; OT2 OD023206; and Community Partners (OT2 OD025277; OT2 OD025315; OT2 OD025337; OT2 OD025276). Results reported are in compliance with the All of Us Data and Statistics Dissemination Policy disallowing disclosure of group counts under 20 to protect participant privacy.

## Availability of Data and Materials

Only data from existing AoU and UKB study cohorts were analyzed and no new source data were collected for this study. Aggregate-level data for the AoU cohort is publicly available at https://databrowser.researchallofus.org/ (Public Tier dataset). Individual-level data for the AoU cohort, available in the Controlled Tier dataset, can be analyzed by approved researchers on the Researcher Workbench. UKB data can be accessed by approved registered users.

## Declaration of Conpeting Interest

AR is an employee of the Complex Disorders Alliance, SG is an SAB member of Our Future Health. JS, GM, SD, KT, KC, MS, CS, AM and SG are employees of PrecisionLife Ltd. SG and GM are shareholders of and have assigned patent rights to PrecisionLife Ltd.

## Author Contributions

SG, JS, GM, SD, CS, and KT contributed to the design of the study. JS designed the reproducibility analyses. GM, SD, KC, MS, and JS performed the analyses described in this manuscript. KC, SD and MS conducted the analyses on the AoU Researcher Workbench. CS, AM, KT and SD analyzed the genes identified and SG, KT, SD and AR helped source the datasets used. All authors contributed to writing the manuscript and consent to publication.

## Data Availability

Only data from existing All of Us and UK Biobank study cohorts were analyzed and no new source data were collected for this study. Aggregate-level data for the All of Us cohort is publicly available at https://databrowser.researchallofus.org/ (Public Tier dataset). Individual-level data for the All of Us cohort, available in the Controlled Tier dataset, can be analyzed by approved researchers on the Researcher Workbench. UK Biobank data can be accessed by approved registered users.

## Abbreviations

AoU: All of Us
FDR: false discovery rate
GWAS: genome-wide association study
IBD: inflammatory bowel disease
PCA: principal component analysis
SNP: single nucleotide polymorphism
UKB: UK Biobank

## Acknowledgenents

We gratefully acknowledge All of Us and UK Biobank participants for their contributions, without whom this research would not have been possible. This work uses data provided by patients and collected by the NHS as part of their care and support, for which we are also grateful. Special thanks to Gert Møller and Claus Erik Jensen, who initially developed the combinatorial analytics methodology, Matthew Pearson and James Kozubek for prior analyses related to endometriosis, and the rest of the PrecisionLife and Complex Disorders Alliance teams for their contributions to supporting the platform and study. Research described in this article has been conducted using data from the All Of Us Research Program Curated Data Repository (CDR) version 8 and UK Biobank (application number 44288) study. We thank the National Institutes of Health’s All of Us Research Program and UK Biobank for making available the participant data examined in this study.

