## Supplemental Data for "Identification and Reproducibility of Novel Combinatorial Genetic Risk Factors for Endometriosis across UK and US Patient Cohorts"

### 1. Supplementary Tables

**Supplementary Table 1.** List of ICD-9 and ICD-10 codes (source concepts) used for endometriosis patient selection in All of Us (AoU)[1].

| AoU Source concept name | ICD-9 code | ICD-10 code |
|---|---|---|
| Endometriosis of ovary | 617.1 | N80.1 |
| Endometriosis of fallopian tube | 617.2 | N80.2 |
| Endometriosis of pelvic peritoneum | 617.3 | N80.3 |
| Endometriosis of rectovaginal septum and vagina | 617.4 | N80.4 |
| Endometriosis of intestine | 617.5 | N80.5 |
| Endometriosis in cutaneous scar/scar of skin | 617.6 | N80.6 |
| Other Endometriosis/Endometriosis of other specified sites | 617.8 | N80.8 |
| Endometriosis, unspecified | 617.9 | N80.9 |
| Endometriosis, site unspecified | | N80.399 |
| Endometriosis of bladder and ureters | | N80.A |
| Endometriosis of bladder, unspecified depth | | N80.A0 |
| Endometriosis of lung | | N80.B2 |
| Endometriosis of the abdomen | | N80.C |
| Endometriosis of the abdomen, unspecified | | N80.C0 |
| Endometriosis of the anterior abdominal wall, unspecified | | N80.C19 |
| Endometriosis of other site of abdomen | | N80.C9 |
| Endometriosis of the pelvic nerves | | N80.D |
| Endometriosis of the pelvic nerves, unspecified | | N80.D0 |

**Supplementary Table 2**. List of conditions or procedures used for exclusion criteria of endometriosis controls using ICD-9 and ICD-10 codes that are also known as AoU Source concepts.

| AoU Source Concepts | ICD-9 codes | ICD-10 codes |
|---|---|---|
| Endometriosis | 617 | N80 |
| Female infertility / Infertility, female | 628 | N97 |
| Dysmenorrhea | 625.3 | N94.4, N94.5, N94.6 |
| Psychogenic dysmenorrhea | 306.52 | |
| Pain in joint, pelvic region and thigh | 719.45 | |
| Pelvic and perineal pain | | R10.2 |
| Pelvic congestion syndrome | 625.5 | |
| Laparoscopy | 54.21 | |

| | |
|---|---|
| Unlisted procedure, laparoscopy, hysteroscopy. | 68.12 |

**Supplementary Table 3.** List of conditions used for exclusion criteria of endometriosis controls using AoU Standard concepts (SNOMED codes) or observation codes.

| **Concept or Code Type** | **Concept or Code name** |
|---|---|
| **Observations codes** | Antenatal care: history of infertility |
| | Infertility care |
| **AoU Standard Concepts** | Acute pelvic pain |
| | Bony pelvic pain |
| | Chronic female pelvic pain syndrome |
| | Chronic pelvic pain of female |
| | Chronic prostatitis |
| | Chronic prostatitis - chronic pelvic pain syndrome |
| | Cyclic pelvic pain |
| | Dysmenorrhea |
| | Endometriosis (clinical) |
| | Endometriosis of lung |
| | Endometriosis of pelvic peritoneum |
| | Endometriosis of rectovaginal septum |
| | Endometriosis of umbilicus |
| | Endometriosis of uterosacral ligament |
| | Female infertility |
| | Female infertility due to diminished ovarian reserve |
| | Female infertility due to ovulatory disorder |
| | Joint pain of pelvic region |
| | Pelvic and perineal pain |
| | Pelvic congestion syndrome |
| | Pelvic girdle pain |
| | Primary infertility |
| | Psychogenic dysmenorrhea |
| | Secondary infertility |
| | Spasmodic dysmenorrhea |
| | Unexplained infertility |
| | Diagnostic laparoscopy |
| | Diagnostic laparoscopy of female pelvis |
| | Laparoscopic lysis of adhesions |
| | Laparoscopic-assisted vaginal hysterectomy |
| | Laparoscopy |
| | Laparoscopy assisted vaginal hysterectomy with bilateral salpingo-oophorectomy |
| | Laparoscopy of fallopian tube |

| | |
|---|---|
| | Laparoscopy with biopsy |
| | Laparoscopy with fulguration of fallopian tube |
| | Laparoscopy with occlusion of fallopian tube by device |
| | Laparoscopy with removal of adnexal structures |
| **Current Procedural Terminology (CPT) codes** | Laparoscopy, surgical, total hysterectomy for resection of malignancy (tumor debulking), with omentectomy including salpingo-oophorectomy, unilateral or bilateral, when performed |
| | Laparoscopy, surgical, with radical hysterectomy, with bilateral total pelvic lymphadenectomy and para-aortic lymph node sampling (biopsy), with removal of tube(s) and ovary(s), if performed |
| | Laparoscopy, surgical; with fulguration of oviducts (with or without transection). |
| | Laparoscopy, surgical; with fulguration or excision of lesions of the ovary, pelvic viscera, or peritoneal surface by any method. |
| | Laparoscopy, surgical; with occlusion of oviducts by device (eg, band, clip, or Falope ring). |
| | Laparoscopy, surgical; with vaginal hysterectomy with or without removal of tube(s), with or without removal of ovary(s) (laparoscopic assisted vaginal hysterectomy). |
| | Unlisted laparoscopy procedure, oviduct, ovary |
| | Unlisted laparoscopy procedure, uterus |
| **Survey codes** | Have you or anyone in your family ever been diagnosed with the following conditions? Think only of the people you are related to by blood. Select all that apply. - Endometriosis |
| | Including yourself, who in your family has had endometriosis? Select all that apply. - Daughter |
| | Including yourself, who in your family has had endometriosis? Select all that apply. - Grandaparent |
| | Including yourself, who in your family has had endometriosis? Select all that apply. - Mother |
| | Including yourself, who in your family has had endometriosis? Select all that apply. - Self |
| | Including yourself, who in your family has had endometriosis? Select all that apply. - Sibling |

**Supplementary Table 4.** Demographic breakdown of cases and controls in the All of Us (AoU) endometriosis cohort. "Other self-reported race" includes American Indian or Alaska Native, Middle Eastern or North African and Native Hawaiian or Other Pacific Islander.

| Category | Value | # Cases (n = 4,134) | % Cases | # Controls (n = 16,536) | % Controls |
|---|---|---|---|---|---|
| **Self-reported race** | White | 2745 | 66.4 | 10722 | 64.84 |
| **Self-reported race** | Black or African American | 506 | 12.24 | 2184 | 13.21 |
| **Self-reported race** | Asian | 92 | 2.23 | 354 | 2.14 |
| **Self-reported race** | Other self-reported race | 47 | 1.13 | 190 | 1.14 |
| **Self-reported race** | More than one population | 246 | 5.95 | 998 | 6.04 |
| **Self-reported race** | None Indicated | 498 | 12.05 | 2088 | 12.63 |
| **Self-reported Ethnicity** | Hispanic or Latino | 629 | 15.22 | 2619 | 15.84 |
| **Self-reported Ethnicity** | Not Hispanic or Latino | 3505 | 84.78 | 13917 | 84.16 |

**Supplementary Table 5.** Principal component analysis (PCA) eigenvalue for AoU endometriosis dataset and self-reported ancestry sub-cohorts. PCs highlighted in bold were selected as covariates in the logistic regression to control for population substructure.

| Principal Component | Eigenvalues -all ancestry | Eigenvalues – self-reported Black / African-American | Eigenvalues – self-reported Hispanic / Latino |
|---|---|---|---|
| **PC1** | **502.17** | **14.67** | **78.71** |
| **PC2** | **246.82** | **2.40** | **23.61** |
| **PC3** | **62.22** | 2.08 | 3.19 |
| **PC4** | **31.23** | 2.05 | 2.80 |
| PC5 | 8.55 | 1.81 | 2.48 |
| PC6 | 7.79 | 1.80 | 2.10 |
| PC7 | 6.54 | 1.78 | 2.00 |
| PC8 | 4.17 | 1.76 | 1.99 |
| PC9 | 3.74 | 1.75 | 1.97 |
| PC10 | 3.64 | 1.74 | 1.95 |
| PC11 | 3.31 | 1.74 | 1.94 |
| PC12 | 3.12 | 1.72 | 1.93 |
| PC13 | 2.69 | 1.70 | 1.91 |
| PC14 | 2.63 | 1.69 | 1.90 |
| PC15 | 2.61 | 1.69 | 1.88 |
| PC16 | 2.59 | 1.68 | 1.88 |
| PC17 | 2.58 | 1.67 | 1.86 |
| PC18 | 2.57 | 1.67 | 1.86 |
| PC19 | 2.56 | 1.65 | 1.85 |
| PC20 | 2.56 | 1.64 | 1.84 |

**Supplementary Table 6.** Comparison of gene mapping of 18 lead meta-GWAS SNPs in disease signatures identified by PrecisionLife platform with their genes associated in Rahmioglu et al. (2023)[2].

| Lead meta-GWAS SNP | Gene(s) associated in meta-GWAS | Gene(s) associated in this study for biological interpretation |
|---|---|---|
| rs507666 | *ABO* | *ABO* |
| rs2967684 | *ACTL9* | |
| rs4540228 | *CD109* | |
| rs10122243 | *CDKN2B-AS1* | |
| rs7334326 | *DLEU1* | *DLEU1* |
| rs5933091 | *FRMD7* | |
| rs3858429 | *FSHB / ARL14EP* | *ARL14EP* |
| rs10090060 | *GDAP1* | *GDAP1* |
| rs11674184 | *GREB1* | *GREB1* |
| rs10860864 | *IGF1* | *IGF1* |
| rs1903068 | *KDR* | |
| rs10828249 | *MLLT10* | |
| rs7907732 | *RNLS* | *RNLS* |
| rs66683298 | *SKAP1* | *SKAP1 / SKAP1-AS1* |
| rs71575922 | *SYNE1* | *SYNE1* |
| rs13441059 | *TEX11* | |
| rs10917151 | *WNT4 / CDC42* | *CDC42* |
| rs55909142 | *7p12.3* | |

**Supplementary Table 7:** Public data sources used for annotating SNPs and genes in the PrecisionLife platform.

| Annotation Type | Data Sources |
|---|---|
| Genomic Context | Ensembl[3], Entrez[4], gnomAD[5], HaploReg[6] |
| Encoded Protein Structure and Function | Uniprot[7], InterPro[8], PDBe-KB[9] |
| SNP-Disease or Trait Association | dbSNP[10], GWAS Catalog[11], PharmGKB[12], Open Targets Genetics[13] |
| Gene-Disease or Trait Association | Open Targets Platform[14,] PHAROS[15], Mouse Genome Informatics[16], Human Phenotype Ontology[17,] OMIM[18], Europe PMC[19] |
| Gene Tissue/Cell Expression | Human Protein Atlas[20], GTEx[21], Expression Atlas[22] |
| Pathways or MoA | Gene Ontology[23], Reactome[24], WikiPathways[25] |
| Drugs and Chemical Compounds | Global Data[26], ChEMBL[27], DrugBank[28], PHAROS[15], PubChem[29] |

**Supplementary Table 8.** Reproducibility statistics in AoU for 35 meta-GWAS SNPs from Rahmioglu et al. (2023)[2].

| SNP ID | Position (GRCh38) | Associated Gene from meta-GWAS | Odds Ratio in All of Us (**bold** = non-reproducing) | *p*-value in All of Us (**bold** = significant with Benjamini-Hochberg FDR correction) |
|---|---|---|---|---|
| rs10917151 | chr1:22096228 | *WNT4/CDC42* | 1.13 | **0.0005** |
| rs12030576 | chr1:115274600 | *NGF* | 1.02 | 0.51 |
| rs2040445 | chr1:169247174 | *SLC19A2* | 1.11 | 0.22 |
| rs2421985 | chr1:172129996 | *DNM3* | 1.02 | 0.52 |
| rs11674184 | chr2:11581409 | *GREB1* | 1.005 | 0.86 |
| rs1430787 | chr2:67641366 | *ETAA1* | 1.07 | 0.02 |
| rs6435157 | chr2:202576501 | *BMPR2* | **0.99** | 0.83 |
| rs1352889 | chr3:49614715 | *BSN* | 1.06 | 0.09 |
| rs1903068 | chr4:55142310 | *KDR* | 1.10 | **0.001** |
| rs2946160 | chr5:158464870 | *EBF1* | 1.02 | 0.48 |
| rs6456259 | chr6:19761487 | *ID4* | 1.04 | 0.56 |
| rs4540228 | chr6:73903344 | *CD109* | 1.08 | **0.003** |
| rs71575922 | chr6:152232879 | *SYNE1* | 1.05 | 0.221 |
| rs11756073 | chr6:170048356 | *FAM120B* | 1.07 | 0.03 |
| rs6970537 | chr7:27177740 | *HOXA10* | 1.05 | 0.04 |
| rs55909142 | chr7:46634176 | *7p12.3* | 1.05 | 0.05 |
| rs10090060 | chr8:74345373 | *GDAP1* | 1.07 | **0.007** |
| rs12549438 | chr8:99138143 | *VPS13B* | 1.02 | 0.55 |
| rs10122243 | chr9:22158925 | *CDKN2B-AS1* | 1.12 | **0.00001** |
| rs507666 | chr9:116708094 | *ASTN2* | 1.02 | 0.48 |
| rs10983311 | chr9:133273983 | *ABO* | 1.05 | 0.14 |
| rs10828249 | chr10:21535798 | *MLLT10* | 1.09 | **0.0006** |
| rs7907732 | chr10:88423007 | *RNLS* | 1.07 | 0.11 |
| rs3858429 | chr11:30322210 | *FSHB/ARL14EP* | 1.07 | 0.049 |
| rs56090796 | chr12:15253294 | *PTPRO* | 1.04 | 0.16 |
| rs3803042 | chr12:53994163 | *HOXC10* | **0.996** | 0.88 |
| rs12320196 | chr12:95251609 | *VEZT* | 1.07 | **0.009** |
| rs10860864 | chr12:102429084 | *IGF1* | 1.001 | 0.97 |
| rs7334326 | chr13:50393596 | *DLEU1* | 1.03 | 0.38 |
| rs66683298 | chr17:48200386 | *SKAP1* | 1.02 | 0.50 |
| rs7214750 | chr17:65907001 | *CEP112* | 1.01 | 0.77 |
| rs2967684 | chr19:8676789 | *ACTL9* | 1.09 | **0.006** |
| rs13441059 | chrX:70889039 | *TEX11* | 1.03 | 0.2955 |
| rs5933091 | chrX:132144079 | *FRMD7* | 1.11 | **0.00007** |
| rs73241342 | chrX:134557708 | *LINC00629* | 1.29 | **0.0003** |

**Supplementary Table 9.** Reproducibility of UKB disease signatures linked to meta-GWAS SNPs and their associated genes from Koller et. al (2026)[30].

| Meta-GWAS gene(s) | Lead 2026 meta-GWAS SNP | # signatures with lead meta-GWAS SNP | Reproducibility rate of meta-GWAS SNP signatures in AoU (OR > 1) | # Signatures assigned to meta-GWAS gene | Reproducibility rate of meta-GWAS gene signatures in AoU (OR > 1) |
|---|---|---|---|---|---|
| *AC007319.1* | rs8176498 | 0 | n/a | 7 | 86% |
| *CCDC26* | rs7828227 | 0 | n/a | 6 | 50% |
| *FOXP1* | rs62246055 | 0 | n/a | 1 | 0% |
| *HMGN1P19* | rs993183 | 0 | n/a | 10 | 60% |
| *HRH1* | rs184700 | 0 | n/a | 2 | 0% |
| *LINC00620* | rs2974396 | 0 | n/a | 6 | 50% |
| *RSPO3* | rs9491697 | 0 | n/a | 1 | 0% |
| *RUNX1* | rs2834747 | 0 | n/a | 2 | 0% |
| *TRPS1* | rs800880 | 0 | n/a | 1 | 100% |

**Supplementary Table 10.** Prior associations with endometriosis among genes identified by PrecisionLife that map to disease signatures with over 9% frequency in All of Us (AoU). This table lists 20 genes, of which 7 are meta-GWAS[2] genes and 16 are associated with endometriosis from Open Targets[14] and Pharos[15].

| **Gene** | **Identified in meta-GWAS** | **Open Targets Evidence** | **Open Targets Gene-Disease Association Score** (using equal weights) | **Pharos Evidence** |
|---|---|---|---|---|
| *ADAM12* | | Differential Expression, Literature | 0.13 | Differential Expression |
| *ARL14EP* | Yes | Genomic association, Literature | 0.53 | |
| *CADM1* | | Literature | 0.05 | |
| *CD34* | | Literature | 0.05 | |
| *CDC42* | Yes | Genomic association, Literature | 0.44 | Genomic association, Literature |
| *CDKN2A* | | Genomic association, Literature | 0.27 | |
| *CDKN2B-AS1* | Yes | Literature | 0.39 | |
| *DSCAM* | | Literature | 0.02 | |
| *FGB* | | Literature | 0.03 | |
| *GLRA3* | | Literature | 0.19 | |
| *GREB1* | Yes | Genomic association, Literature | 0.55 | Genomic association, Literature |
| *IGF1* | Yes | Genomic association, Literature | 0.53 | Genomic association, Literature |

| | | | | |
|---|---|---|---|---|
| *IGF2BP3* | | Genomic association, Literature | 0.10 | Genomic association |
| *IL17RE* | | Literature | 0.14 | |
| *NEDD4L* | | Literature | 0.01 | Literature |
| *POP4* | | Genomic association, Literature | 0.10 | |
| *PTPRD* | | Literature | 0.06 | |
| *RRM2* | | Differential Expression, Literature | 0.13 | Differential Expression |
| *SKAP1* | Yes | Genomic association, Literature | 0.21 | |
| *SYNE1* | Yes | Genomic association, Literature | 0.41 | Genomic association |

**Supplementary Table 11.** Association of endometriosis-associated genes with infertility phenotypes in All of Us. Statistical significance was assessed using Fisher's exact test. Odds ratios (ORs) and corresponding Fisher's exact test *p*-values are shown for each gene analyzed. Only associations with >=20 individuals with infertility phenotype have been reported.

| Gene | Phenotype | Frequency cutoff of Reproduced Signatures in AoU | Odds Ratio | *p*-value before multiple testing correction |
|---|---|---|---|---|
| *AL049792.1* | Female infertility (AoU Standard Concept) | 1% | 1.791 | 0.021 |
| *AL445253.1* | Female infertility (AoU Standard Concept) | 1% | 1.227 | 0.035 |
| *ATG16L1* | Female infertility (AoU Standard Concept) | 1% | 1.403 | 0.014 |
| *CACNA2D3* | Female infertility (AoU Standard Concept) | 1% | 1.372 | 0.015 |
| *CCDC3* | Female infertility (AoU Standard Concept) | 1% | 1.825 | 0.002 |
| *CCSER1* | Female infertility (AoU Standard Concept) | 1% | 1.289 | 0.018 |
| *DEAF1* | Female infertility (AoU Standard Concept) | 1% | 1.467 | 0.031 |
| *DNAH11* | Female infertility (AoU Standard Concept) | 1% | 1.388 | 0.031 |
| *ETV1* | Female infertility (AoU Standard Concept) | 1% | 1.539 | 0.033 |
| *GPR35* | Female infertility (AoU Standard Concept) | 1% | 1.539 | 0.033 |
| *KCNB2* | Female infertility (AoU Standard Concept) | 1% | 2.011 | 0.006 |
| *KIF12* | Female infertility (AoU Standard Concept) | 1% | 1.382 | 0.029 |
| *MAML3* | Female infertility (AoU Standard Concept) | 1% | 1.814 | 0.014 |
| *NINJ1* | Female infertility (AoU Standard Concept) | 1% | 1.791 | 0.021 |
| *POLR3KP1* | Female infertility (AoU Standard Concept) | 1% | 1.939 | 0.003 |
| *PRKCA* | Female infertility (AoU Standard Concept) | 1% | 1.406 | 0.019 |
| *PRKG1-AS1* | Female infertility (AoU Standard Concept) | 1% | 2.011 | 0.006 |
| *RSRC1* | Female infertility (AoU Standard Concept) | 1% | 1.901 | 0.004 |
| *SCN2A* | Female infertility (AoU Standard Concept) | 1% | 1.939 | 0.003 |
| *TMEM179* | Female infertility (AoU Standard Concept) | 1% | 2.011 | 0.006 |
| *DEC1* | Female infertility (AoU Standard Concept) | 9% | 1.270 | 0.038 |
| *LRP1B* | Female infertility (AoU Standard Concept) | 9% | 1.448 | 0.002 |
| *AC007342.4* | Female infertility (ICD10 N97.x) | 1% | 1.712 | 0.025 |
| *AGO4* | Female infertility (ICD10 N97.x) | 1% | 1.461 | 0.041 |

| | | | | |
|---|---|---|---|---|
| *AL445253.1* | Female infertility (ICD10 N97) | 1% | 1.290 | 0.046 |
| *ATG16L1* | Female infertility (ICD10 N97) | 1% | 1.668 | 0.003 |
| *CCSER1* | Female infertility (ICD10 N97) | 1% | 1.348 | 0.030 |
| *ETV1* | Female infertility (ICD10 N97) | 1% | 2.048 | 0.003 |
| *GPR35* | Female infertility (ICD10 N97) | 1% | 2.048 | 0.003 |
| *LARP7P3* | Female infertility (ICD10 N97) | 1% | 1.809 | 0.024 |
| *NIPAL2* | Female infertility (ICD10 N97) | 1% | 1.650 | 0.043 |
| *NPNT* | Female infertility (ICD10 N97) | 1% | 1.724 | 0.030 |
| *USP9X* | Female infertility (ICD10 N97) | 9% | 1.450 | 0.044 |
| *WNT9A* | Female infertility (ICD10 N97) | 9% | 1.450 | 0.044 |
| *GLRA3* | Assistive reproductive technology | 1% | 1.967 | 0.013 |
| *PWWP3B* | Assistive reproductive technology | 1% | 1.693 | 0.040 |
| *Z82186.1* | Assistive reproductive technology | 1% | 1.725 | 0.048 |

**Supplementary Table 12.** List of infertility phenotype codes in All of Us (AoU) used in this study.

| **Phenotype** | **Total no. of Individuals in AoU** | **No. of Endometriosis cases in AoU (% of 4134 cases)** |
|---|---|---|
| Female infertility (AoU Standard Concept, SNOMED Code: 6738008) | 5949 | 557 (13.47%) |
| Female infertility ICD10 N97 | 3600 | 302 (7.30%) |
| Assistive Reproductive Technology (CPT codes : 58970, 58974, 58976, 89250-89259) | 613 | 64 (1.55%) |
| Any evidence of Infertility from the above codes | 6029 | 557 (13.47%) |

### 2. Supplementary Figures

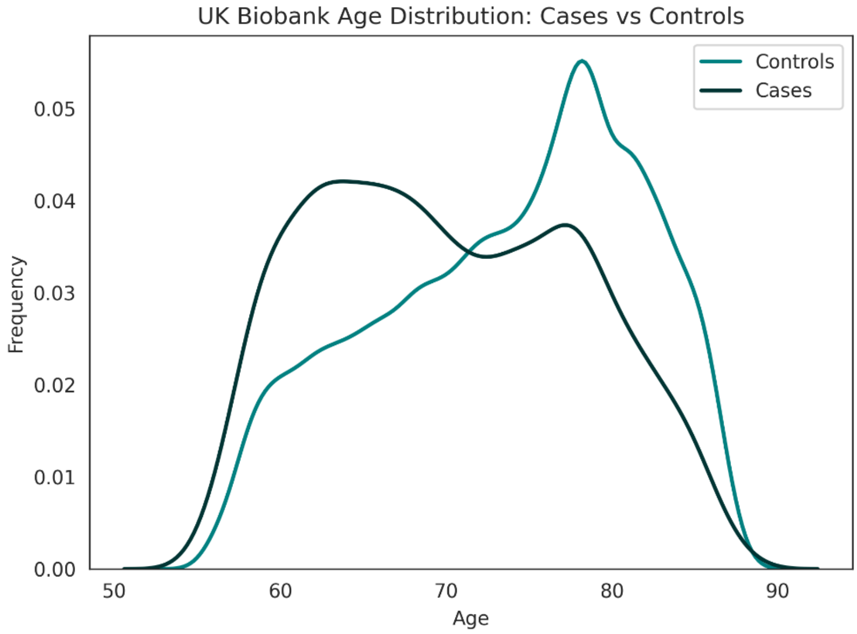

**Supplementary Figure 1.** Distribution of ages for endometriosis cases and controls in UK Biobank cohort. Age calculated as 2025 - Year of Birth.

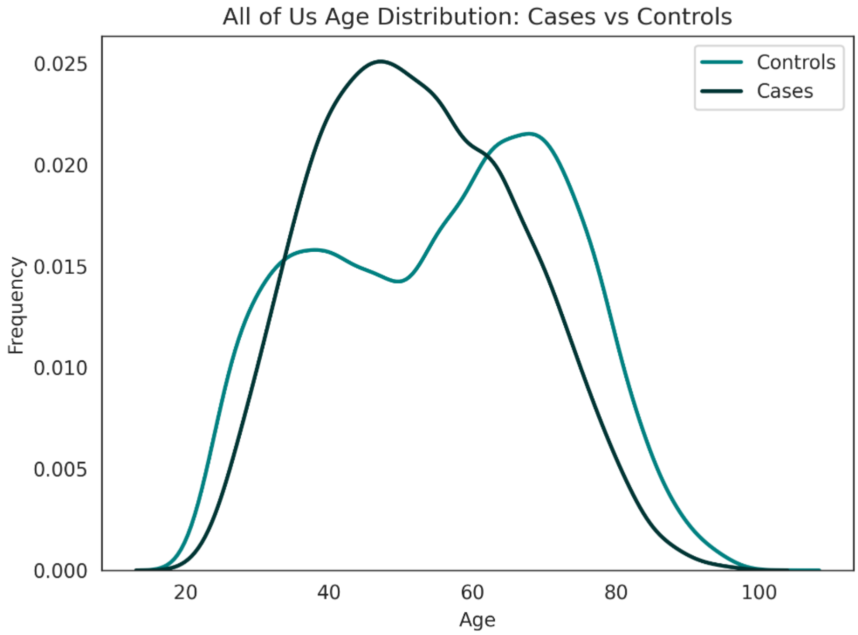

**Supplementary Figure 2.** Distribution of ages for endometriosis cases and controls in All of Us validation cohort. Age calculated as 2025 - Year of Birth.

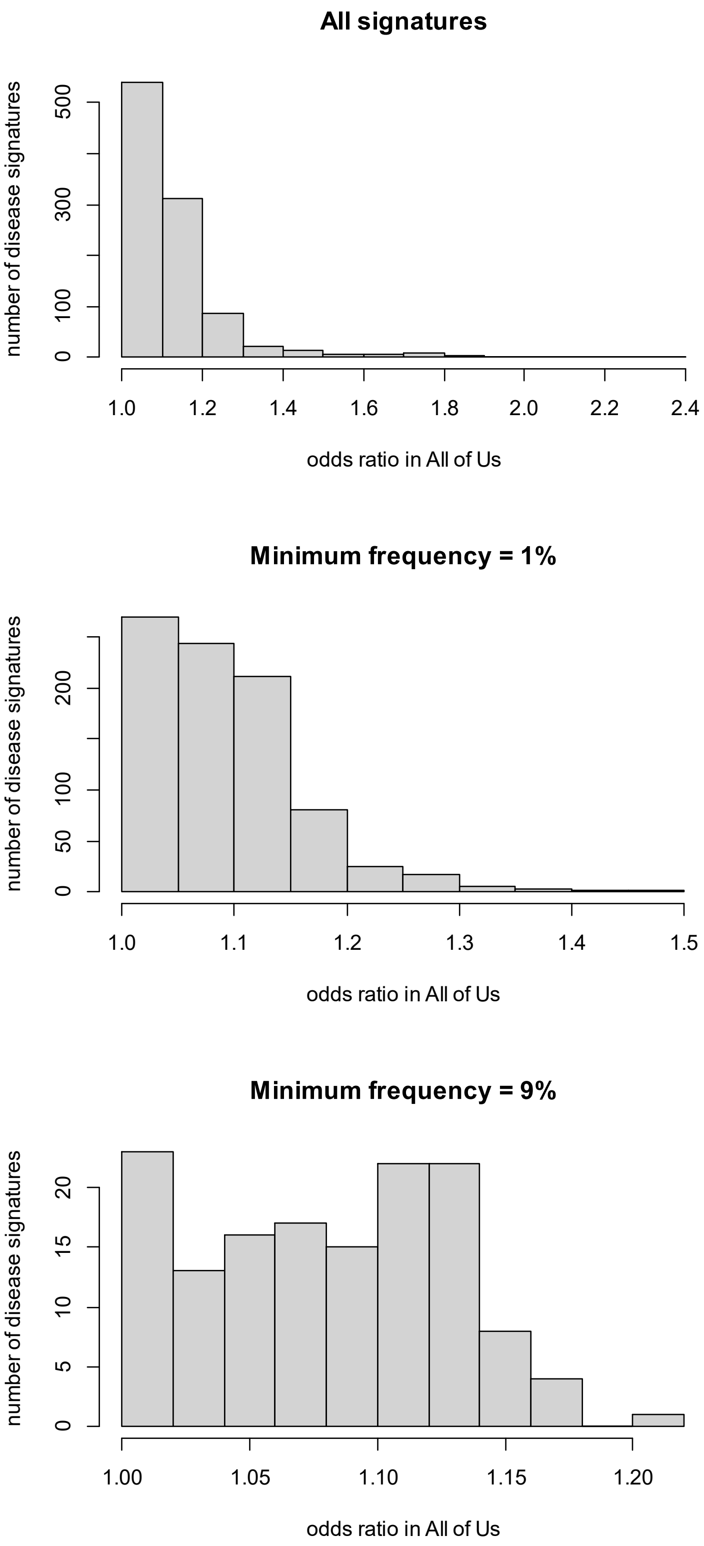

**Supplementary Figure 3.** Odds ratios of reproducing signatures in All of Us. Top: all disease signatures. Middle: disease signatures with frequency > 1%. Bottom disease signatures with frequency > 9%

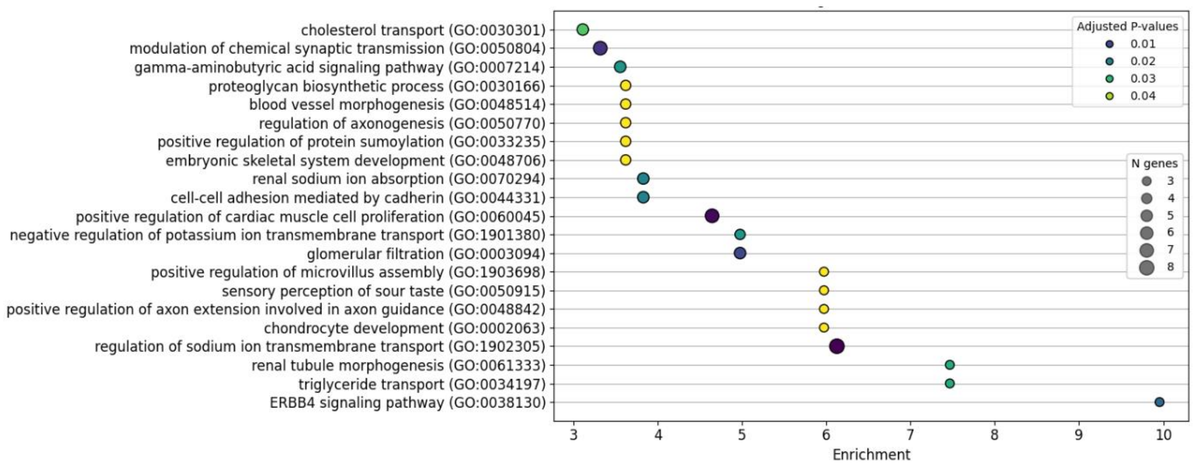

(a)

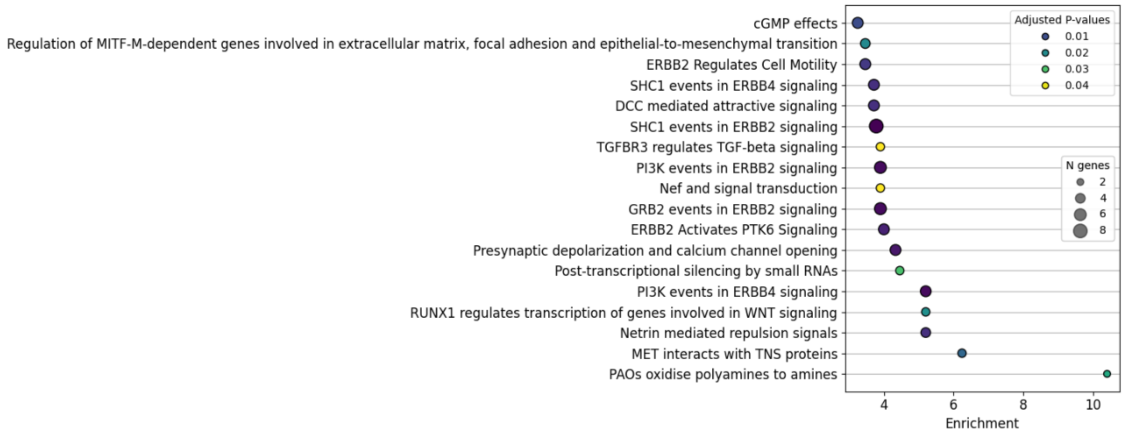

(b)

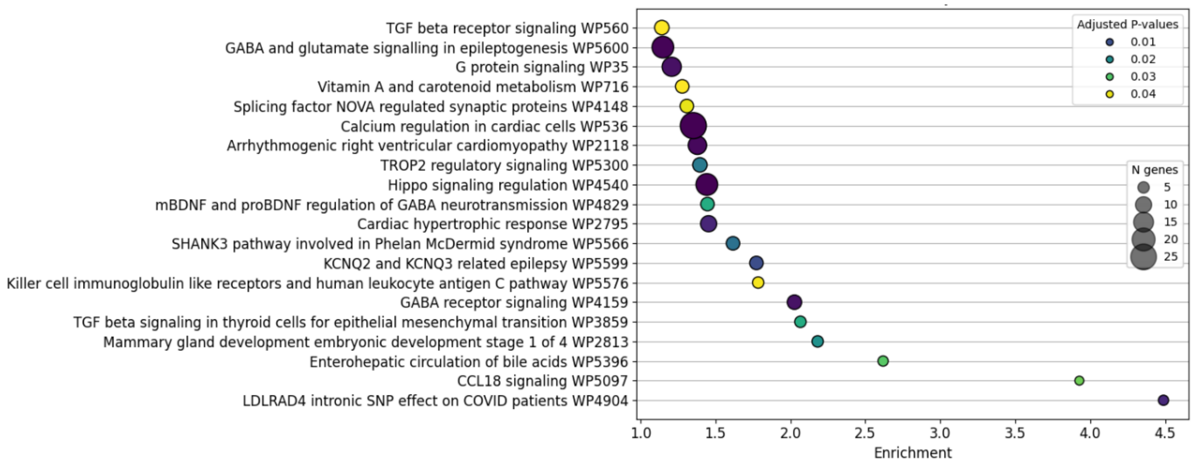

(c)

**Supplementary Figure 4.** Gene set enrichment plot for significant (adjusted *p*-values < 0.05) biological processes using (a) Gene Ontology, (b) Reactome and (c) WikiPathways for genes mapped to all combinatorial disease signatures (n=1709). Each dot in the plot represents an enriched gene set representing a biological process or pathway. The y-axis lists pathway names, and the x-axis indicates the enrichment score, reflecting the strength of over-representation of the gene set in the input list. Dot size corresponds to the number of genes associated with each gene set and the dot color denotes the adjusted p-value, with darker colors indicating higher statistical significance.

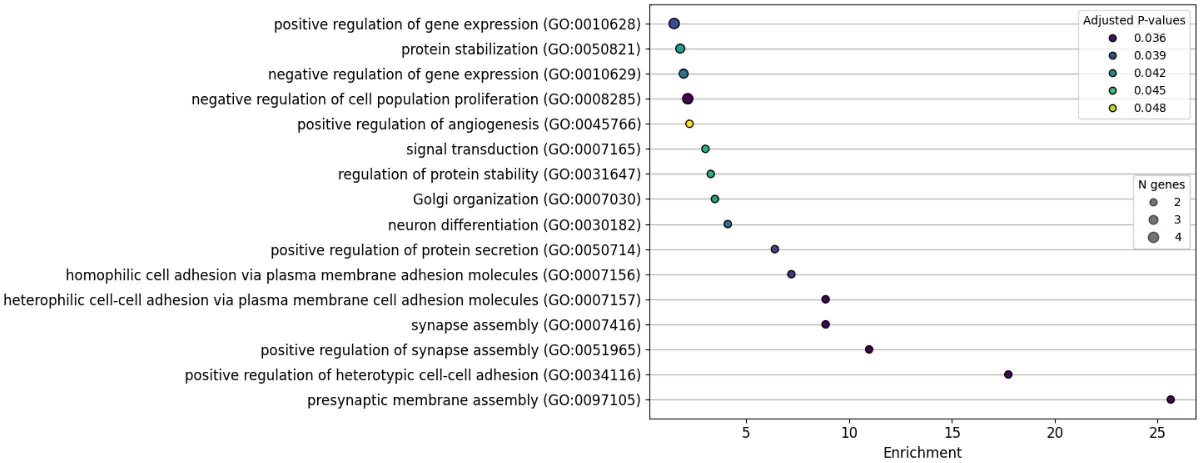

**Supplementary Figure 5.** Gene set enrichment plot for significant (adjusted *p*-values < 0.05) biological processes (Gene Ontology) for genes mapped to disease signatures with frequency > 9% (n=141) in All of Us. The enriched GO biological process terms are listed in Extended Table 5. Reactome and WikiPathways did not generate any significant results. Each dot in the plot represents an enriched gene set representing a biological process or pathway. The y-axis lists pathway names, and the x-axis indicates the enrichment score, reflecting the strength of over-representation of the gene set in the input list. Dot size corresponds to the number of genes associated with each gene set and the dot color denotes the adjusted p-value, with darker colors indicating higher statistical significance.

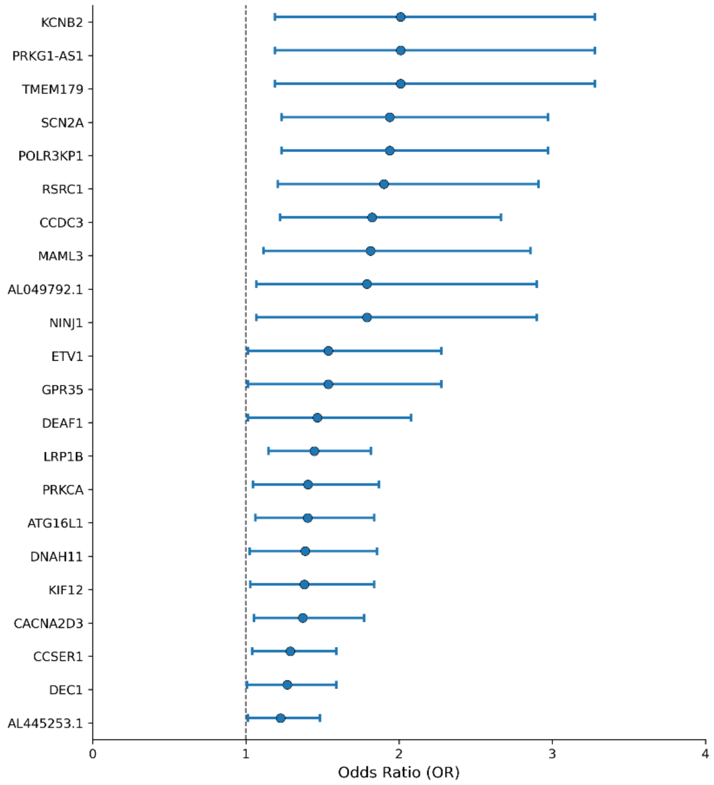

(a) Female infertility AoU Standard Concept (SNOMED Code: 6738008)

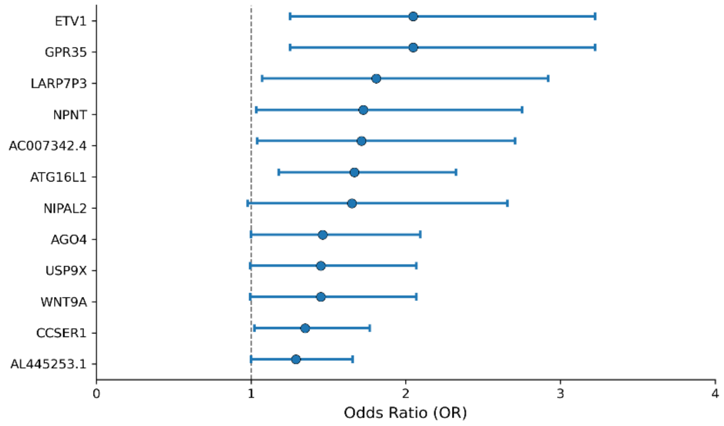

(b) Female infertility ICD10 N97

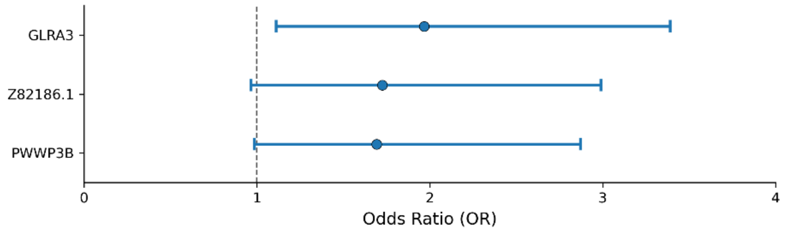

(c) Assistive Reproductive Technology

**Supplementary Figure 6.** Association between endometriosis-associated genes and infertility phenotypes in All of Us with odds ratio > 1 and *p*-value < 0.05 before multiple-testing correction. Forest plot showing odds ratios (ORs) and 95% confidence intervals (CIs) for the association of each gene (listed on the y-axis) with (a) female infertility AoU Standard Concept (SNOMED Code: 6738008), (b) female infertility ICD10 N97 and (c) assistive reproductive technology. Points represent OR estimates and horizontal lines indicate 95% CIs. The dashed vertical line denotes OR = 1.
